# Population scale proteomics enables adaptive digital twin modelling in sepsis

**DOI:** 10.1101/2024.03.20.24304575

**Authors:** Aaron M. Scott, Lisa Mellhammar, Erik Malmström, Axel Goch Gustafsson, Anahita Bakochi, Marc Isaksson, Tirthankar Mohanty, Louise Thelaus, Fredrik Kahn, Lars Malmström, Johan Malmström, Adam Linder

## Abstract

Sepsis is one of the leading causes of mortality in the world. Currently, the heterogeneity of sepsis makes it challenging to stratify patients on admission to the emergency department, hindering the implementation of precision medicine. Here, we acquired clinical parameters and blood plasma proteome profiles for 3009 patient samples to develop a framework for digital twin-based modelling in sepsis. Already on admission to the emergency department, the model can accurately diagnose sepsis and infection, while providing prognostic predictions for persistent organ dysfunction, mortality, and intensive care unit admission. Furthermore, the model can suggest therapeutically actionable pathways based on patient-unique proteome trajectories, determine the infection loci and pathogen type, and identify which patients may benefit from vasopressor treatment at the time-of-admission to the emergency department. The framework has the potential to advance precision medicine in sepsis and provides a generalizable approach for data-driven disease modeling and clinical decision support.

## Introduction

Sepsis is a clinical syndrome that has been re-defined as life-threatening organ dysfunction caused by a dysregulated host response to infection (sepsis-3)^1^. Despite its clinical impact, and over 100 clinical intervention studies^2^, the mortality of sepsis remains high and the definition imprecise^2^. Sepsis patients are a heterogeneous population in terms of age, underlying comorbidities, infecting pathogen, and the infection foci. In addition, sepsis patients present complex pathophysiology, including concomitant inflammation and immunosuppression with rapid disease progression, further augmenting the heterogeneity of the syndrome. Currently, the severity of organ dysfunction in sepsis is measured by the Sequential Organ Failure Assessment (SOFA) scoring system and the standardized treatment strategy for sepsis includes early broad-spectrum antibiotic therapy, source control, and organ supportive care. Modulating the host response to infection has been proposed as a treatment strategy without sufficient efficacy, although reports indicate that such treatments may be beneficial for some patients^2^. Consequently, developing improved stratification strategies has the potential to catalyze the introduction of precision medicine in sepsis where treatments are given to enriched groups of responding patients.

Previous attempts to stratify sepsis into more homogeneous subgroups have focused on subdividing patients based on clinical or molecular data^3–9^. Optimally, subphenotypes are defined by a minimal combination of features to facilitate clinical implementation, and such approaches have shown to be useful in the treatment and definition of other diseases^10–14^. In sepsis, subphenotyping strategies have facilitated the discovery of biologically similar groups of patients with different clinical outcomes and predicted treatment responses ^5,7–9,15,16^. Unfortunately, the results of subphenotyping studies in sepsis are not always comparable, and sometimes contradictory, as different studies may identify different phenotypic aspects of sepsis ^10,17^. To combat this issue, other recent efforts focus on combining multiple subphenotype definitions using consensus clustering to give a more precise estimate of outcome compared to single subtypes ^7,8,17^. Recent studies have also shown that heterogeneity within subtypes can lead to patient-subtype assignment changes over short intervals and varying effects of early goal-directed therapy, where risk-benefit variability within a subtype can lead to harmful outcomes for certain patients^18–21^. To make subphenotypes in sepsis more feasible, a shift from broad subtypes to those focused on specific treatable traits could maximize clinical utility for patients in the future^22,23^. Overall, due to the diverse results of subphenotyping in sepsis, a more personalized approach for patient stratification could supplement existing efforts and accelerate the translation of proposed patient care frameworks in sepsis to the clinic.

Digital twin (DT) modeling has emerged as a promising framework for understanding and predicting complex biological and clinical processes. By creating virtual representations of individual patients or diseases, digital twins enable dynamic simulation of disease progression and treatment responses that integrate molecular, clinical, and temporal data ^24–27^ . This approach moves beyond subphenotype models to capture patient-specific variability, allowing for real-time prediction, continuous model refinement, and personalized therapeutic guidance. The concept has been empowered by advances in generative artificial intelligence and has contributed to disease understanding, biomarker discovery, and drug development^28–32^. There are many different flavors of digital twins in medicine, and no set upon standard approach. Some efforts focus on combining multimodal data to build in-silico representations of individual patients to predict outcome in cancer ^27,33^, or build computational models of the heart to study cardiovascular disease^34^. Other methods focus on modelling heterogeneous diseases through the construction of patient similarity networks^35–37^ . Regardless of the specific approach, the general concept of a digital twin in medicine revolves around building computational models that mimic new patients or diseases at different levels to provide diagnostic or prognostic predictions and identify potential treatment options.

To date, aside from a pioneering prospective clinical observation study involving 29 patients in the intensive care unit (ICU)^38^, digital twin modeling has not yet been applied to sepsis. Implementing digital twin modeling in this context requires large, high-quality datasets that integrate diverse data types collected under standardized conditions. Furthermore, constructing accurate and generalizable digital twins necessitates specialized computational solutions that embrace the heterogeneous nature of sepsis.

## Results

In this study, we present an interpretable framework for adaptive digital twin modeling in sepsis as a complementary approach for patient stratification at the time-of-admission. The framework accommodates both time-of-admission clinical parameters and plasma proteomic profiles (**Figure 1a**) to generate virtual representations, or digital twins, of the pathological landscape of sepsis. These digital twins are built using diverse population scale cohorts, composed of varying etiologies and prognostic outcomes, by connecting patients to their nearest neighbors to form an extensive graph structure. Due to the scale of the data included, this graph structure provides a comprehensive overview of the progression and inherent heterogeneity of sepsis. On admission to the emergency department (ED), new patients can be integrated with their nearest neighbors in the model to diagnose sepsis and infection, prognosticate future outcomes, and predict patient specific treatment. Beyond traditional stratification, this model can also be used to dynamically track molecular trajectories through disease progression to suggest potential personalized treatment options for individual patients (**Extended Data Figure 2a**).

**Figure 1.**
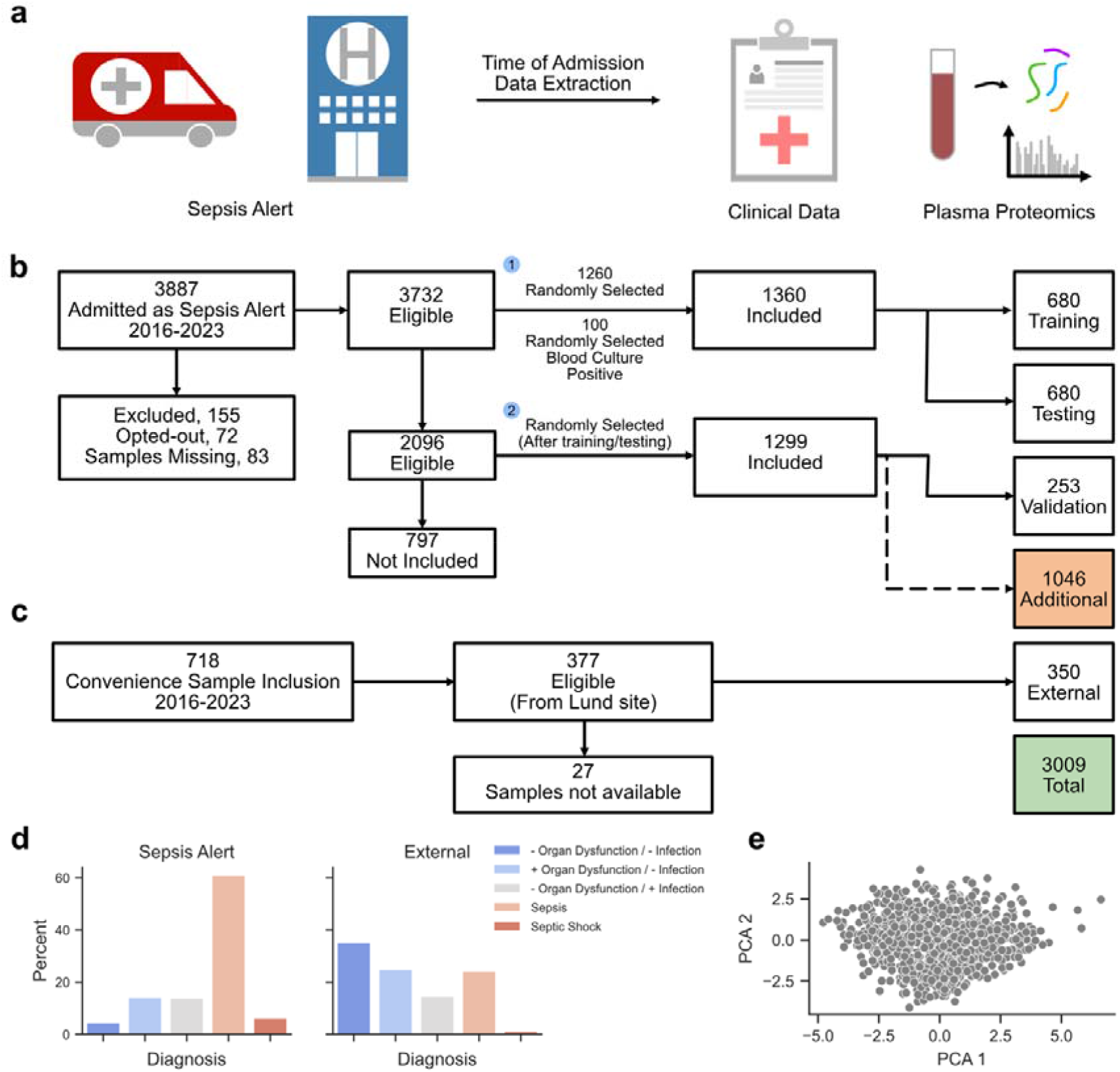
Study overview and patient selection. **a)** A schematic representation of the sample collection process. Briefly, patients were selected on arrival to the emergency department if they were suspected of sepsis and blood samples were taken. Clinical data was extracted for each of these suspected sepsis cases and the plasma proteome of the blood samples were analyzed using mass spectrometry-based proteomics. **b)** An overview of the sepsis alert study cohort used in the study. Of the 3732 patients, 1360 were included in the main study cohort. This was split into the training (n=680) and testing (n=680) cohorts, with 253 additional patients selected later from the eligible patients as the validation cohort. An additional cohort (n=1046) was selected from the eligible patients with partial clinical data extracted from electronic health records **c)** An overview of the patients admitted and included in the external cohort. Of 718 eligible patients, 350 were included to establish the external cohort. **d)** A bar plot visualizing the distributions of sepsis diagnosis groups in the training, testing, validation (Sepsis Alert), and external cohorts. The colors are based on the infection groups: no infection + no organ dysfunction, no infection + organ dysfunction, infection + no organ dysfunction, sepsis (infection + organ dysfunction), and septic shock. **e)** A scatter plot of 2 principal components from principal component analysis (PCA) using 13 common time-of-admission clinical parameters.

In total, 3009 unique samples from two separate study cohorts with different inclusion criteria were included to construct this framework (**Figure 1b-c**). From the first cohort, 3887 patients with suspected infection and the highest triage level, referred to as sepsis alert patients^39^, were admitted to the emergency department (ED) during the study period, with 3732 patients eligible for inclusion (**Figure 1b**). From this cohort, a total of 2670 patients were selected and subdivided into four groups referred to as the training (n=680), testing (n=680), validation (n=253) and additional (n=1046) cohorts. The training, testing, and validation groups retained the ratios of defined diagnosis groups as observed among the 3887 patients in the whole sepsis alert cohort. From the second study cohort, 350 patients were selected from outside the sepsis alert study cohort with different inclusion criteria to generate the external cohort (**Figure 1b**).

Within these cohorts (**Figure 1b**), the following 5 diagnosis groups were defined: no infection and no organ dysfunction (1), no infection with organ dysfunction (2), infection without organ dysfunction (3), infection with organ dysfunction (sepsis-3) (4), and septic shock (5). These groups were based on an increase in SOFA score and the presence of infection, determined using the Linder-Mellhammar Criteria for Infection (LMCI)^40^. The external cohort contained a different distribution of diagnosis groups than the sepsis alert cohort with proportionally fewer sepsis and septic shock patients (**Figure 1d**). For each of the 3009 patients, blood samples were collected at time-of-admission, and the plasma proteome was analyzed using mass spectrometry-based proteomics. For all samples, up to 71 clinical parameters were extracted to create a detailed multi-modal database that incorporates patient outcome and presenting clinical values with their accompanying blood proteome measurements. Overall, this resulted in 5 separate cohorts referred to from here as the training, testing, validation, external, and additional cohorts. Clinical characteristics and demographics for the cohorts are available in **Supplementary Tables T1a-c**.

### Subphenotypes consist of diverse patient outcomes

Initially, we sought to determine if our cohort could be partitioned into clinically distinct subphenotypes. Consistent with previous observations, the patients included in this study exhibited substantial heterogeneity, with no clear clustering structure (**Figure 1e**)^5,18,19^ . To improve statistical power, we combined the training and testing cohorts and used the same approach as state-of-the-art sepsis subphenotyping strategies to determine an optimal patient grouping based on time-of-admission data ^5,7,8^ . As no clear clustering structure was observed (**Extended Data Figure 1a**), we applied a comprehensive consensus clustering method, using k-means, to partition the patients into optimally defined (k=4) groups (**Extended Data Figure 1b-e**). This procedure yielded 4 spatially distinct clusters (**Extended Data Figure 1f**), with distinct clinical characteristics (**Extended Data Figure 1g**). However, the patient outcomes within each cluster were distributed unevenly, forming gradients in which higher values of 30-day-mortality tended to be located at the periphery of each cluster (**Extended Data Figure 1h**). Principle component analysis (PCA) further confirmed a linear association between spatial location and 30-day-mortality (**Extended Data Figure 1i**), quantifying the subphenotype gradients. Additionally, many samples were more similar to neighboring clusters than to their assigned ones, suggesting incorrect cluster assignment (**Extended Data Figure 1j**). Thus, while patients were assigned to clinically distinct subphenotypes with differing average outcomes, these outcomes were distributed unevenly throughout each subphenotype with inconsistent patient-cluster assignment. This is consistent with recent findings where early goal-directed therapy in sepsis subtypes has varying effects for patients within the same subtype^18,19^ . These findings suggest that alternative approaches, such as grouping patients with their most immediately similar neighbors, may complement subphenotyping to address the issue of intra-subphenotype gradients, particularly when guiding individualized treatment options.

### Digital twin models prognosticate future outcome

Based on the results from the subphenotyping analysis, we suggest that the patients included in our study could be organized in a nearest-neighbor graph, grouping patients with similar clinical presentations and outcomes together (**Figure 2a**). Since our cohort included a wide distribution of patients suspected of sepsis at hospital admission, this nearest neighbor graph provides a digital representation of the progression of sepsis, constituting a syndrome level sepsis digital twin model. This model could then be used to diagnose and prognose outcome for new patients at the time-of-admission to the hospital as well as model individual patient trajectories (**Extended Data Figure 2a**).

**Figure 2.**
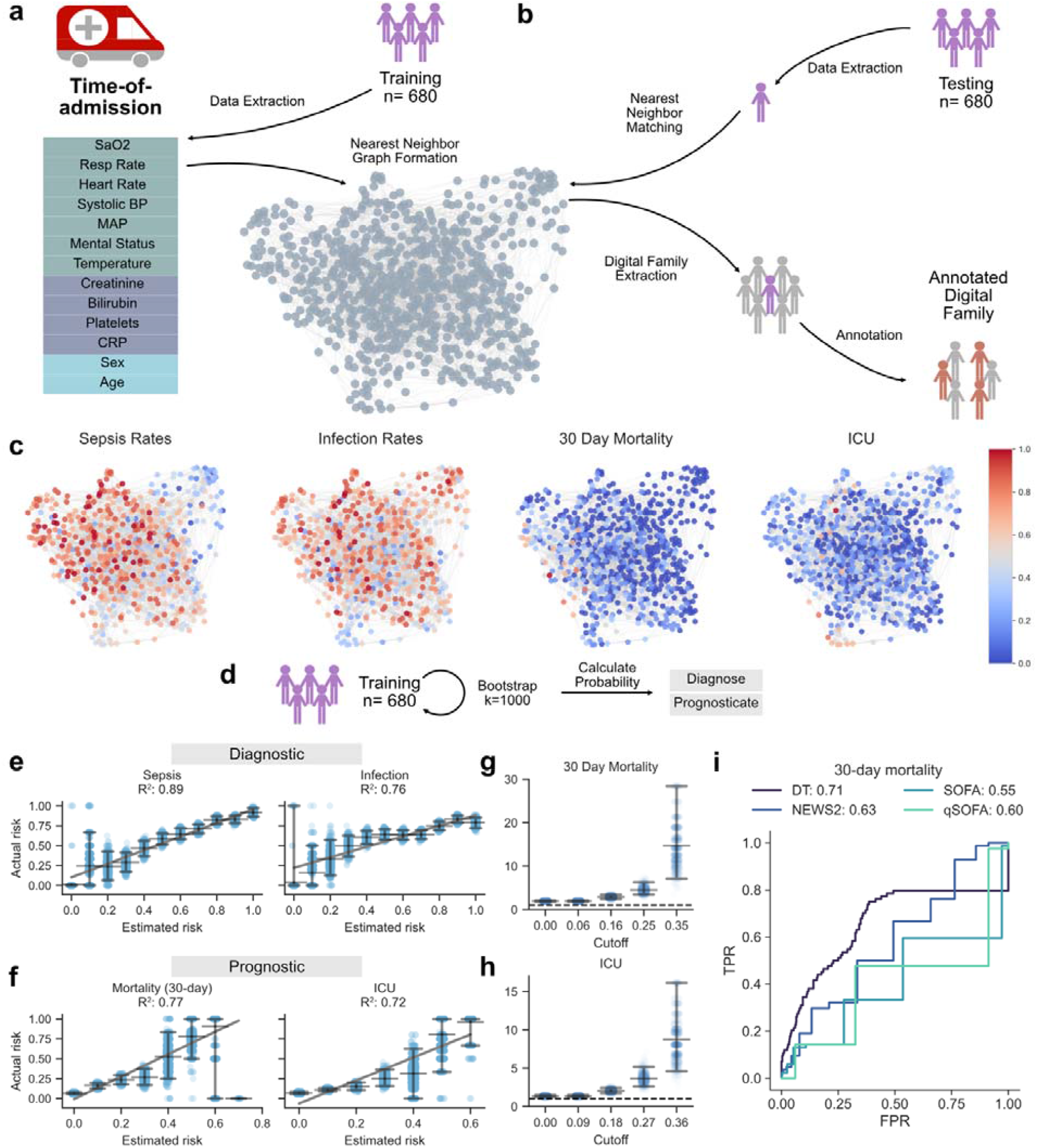
Clinical digital twin models prognosticate future outcomes of suspected sepsis patients with time-of-admission data. **a)** Visualization of the workflow for extracting time-of-admission clinical parameters from patients suspected of sepsis on arrival to the hospital and creation of the digital twin model. Patients arrive at the hospital, are identified as a sepsis alert, and clinical data associated with these patients is extracted, imputed, and normalized. The 13 time-of-admission clinical parameters are then used to build a digital twin model for all 680 patients in the training cohort. **b)** Visualization of the workflow for incorporating new patients into the digital twin model and calculating the estimated probability of future clinical outcomes. Each patient from the testing cohort is treated as a new patient and the 13 time-of-admission clinical parameters are used to integrate the new patient into the digital twin model based on their 10 nearest neighbors. Using these nearest neighbors, risk scores are determined by calculating the frequency of occurrence of a chosen binary clinical outcome. **c)** To evaluate the performance of the digital family model to diagnose and prognose future outcomes, we calculated the estimated risk of sepsis, infection, persistent organ dysfunction (an increase of SOFA scores >= 2 on day 2 and day 3 in the hospital), 30-day mortality, and intensive care unit admission and compared these risk scores to the actual rates in the testing cohort. **d)** 1000 bootstrapping iterations were used to train the model to create prediction intervals for error estimation. **e)** The estimated vs. actual rates for the diagnosis of sepsis and infection as well as squared Pearson correlation coefficients (R^2^). 95% prediction intervals are plotted as well as the background data used to calculate each regression, with a slight jitter added for each estimated rate to visualize the spread of the data. **f)** The estimated vs. actual rates for the prognosis of persistent organ dysfunction, 30-day mortality, and ICU admission as well as squared Pearson correlation coefficients (R^2^). 95% prediction intervals are plotted as well as the background data used to calculate each regression, with a slight jitter added for each estimated rate to visualize the spread of the data. **g-h)** Likelihood ratios for the prognostic predictions of the model at increasing risk levels calculated by the digital twin model and 95% prediction intervals. The cutoffs were determined using the median risk scores with increases based on the incremental median absolute deviation. **g)** Positive likelihood ratios for 30-day mortality **h)** Positive likelihood ratios for ICU admission **i)** A comparison of the CDTM to predict 30-day mortality compared to the NEWS2, SOFA, and qSOFA clinical scores calculated at the time-of-admission. TPR is the true positive rate on the y-axis, and FPR is the false positive rate on the x-axis. The ROC-AUC values are indicated above the figure.

To first determine if the sepsis digital twin model can diagnose as well as prognose future clinical outcomes, we focused on clinical parameters that were available at the time-of-admission and used in routine clinical practice. We extracted 13 common clinical parameters including heart rate, blood pressure, and mental status along with laboratory values such as creatine, bilirubin, and C-reactive protein (CRP) to build a clinical digital twin model (CDTM) from the training cohort (n=680) (**Figure 2a**). To optimize the number of samples used to construct the nearest-neighbor network, we analyzed the intra-neighborhood (family) distance, variation, and diagnostic precision at different numbers of neighbors (k) (**Extended Data Figure 2b-d**) and chose *k*=10 as the number of neighbors to build the digital twin model. The CDTM provides diagnostic potential by aggregating patients together that have similar sepsis and infection diagnoses, and prognostic potential by aggregating patients together that have similar rates of 30-day mortality, and intensive care unit (ICU) admission (**Figure 2c**). In **Figure 2**, each node in the projected network represents a patient in the training data, and the color represents the rate of the diagnostic or prognostic outcome within the 10-nearest neighbors for each patient. This effectively creates zones within the network, where patients with similar clinical outcomes are located close to each other. Based on this concept, when new patients are appended to the digital twin model, diagnostic and prognostic predictions can be made using their nearest neighbors in the network (**Figure 2b**).

To evaluate the performance of CDTM, we used the testing cohort (n=680) as validation to find and extract the nearest neighbors, or digital families, for each new patient, from the sepsis digital twin model (**Figure 2b**). In a clinical setting, it is important that models are highly calibrated when making predictions for individual patients, meaning that predicted risk reflects the observed risk in a population at different levels^41^. A well calibrated model will minimize instances of over, or under, estimation of severe disease, which is essential in the context of sepsis. Based on the frequency of occurrence in the extracted digital family, we calculated the actual rate among all patients in the testing cohort (n=680) and compared to the estimated risk calculated using 1000 bootstrapping iterations from the digital twin model for each clinical outcome to evaluate the calibration of the CDTM (**Figure 2d**). Overall, we observed a high calibration between estimated rate and actual rate for the diagnosis of sepsis (R^2^ = 0.89) and infection (R^2^ = 0.76) (**Figure 2e**), and for the prognoses of 30-day mortality (R^2^ = 0.77), and ICU admission (R^2^ = 0.72) (**Figure 2f**). In this way, the digital twin model provides insight into the clinical environment for new patients in the testing cohort.

To ensure that patients with a possible severe outcome are detected among critically ill patients suspected of sepsis on admission to the emergency department (**Figure 1b**), model sensitivity is essential. Thus, sensitivity, or the true positive rate, was chosen as an initial focus to evaluate our digital twin model. For the prognostic predictions, the CDTM had a sensitivity of 0.80 and 0.89 for 30-day mortality and ICU admission respectively in the testing cohort (n=680). To further demonstrate and highlight the calibration of the model, positive likelihood ratios were also used to evaluate the confidence of positive predictions at increasing risk levels. As the positive likelihood ratio is defined as *sensitivity / (1 – specificity)*, an observed increase in positive likelihood ratios at higher risk thresholds indicates a substantial increase in specificity. For the CDTM, increased prognostic risk is statistically associated with more than 10-times higher likelihood of prognostic outcome based on positive likelihood ratios, indicating a substantial increase in specificity at higher risk levels (**Figure 2g-h**). These metrics are used to evaluate the performance of digital twin models in different contexts throughout our study.

We also compared the CDTM to 3 different clinical risk scores calculated on admission to the emergency department. Our model outperformed all risk scores when prognosing 30-day mortality and was notably superior in areas of clinically relevant false positive rates (<0.20) (**Figure 2i**). These results demonstrate that the digital twin model can accurately diagnose patients in addition to prognose future clinical outcomes with 13 common parameters that are routinely available in clinical practice at the time-of-admission.

### DT models uncover hidden patient similarities

Although the CDTM can stratify patients into different clinical risk groups of diagnostic and prognostic value, it only provides minimal molecular information about individual patient phenotypes. To uncover molecular signatures for patients associated with different sepsis diagnostic risk groups, we incorporated plasma proteomic profiles, to identify molecular changes associated with the low-risk (≤ 0.5) and high-risk (≥0.8) sepsis groups calculated by the CDTM (**Figure 2e** and **Figure 3a** and **Extended Data Figure 3**). Differential abundance analysis uncovered 139 statistically significant proteins between the low-risk and high-risk groups (**Extended Data Figure 3c**), regardless of their clinical diagnosis (**Extended Data Figure 3b**). Gene Ontology (GO) enrichment analysis of the statistically significant protein abundances revealed biological pathways associated with blood coagulation and platelet aggregation in positively enriched proteins (**Extended Data Figure 3d**) and lipid metabolism and transport in negatively enriched proteins (**Extended Data Figure 3e**) in the high-risk group. This indicates a strong biological signal is associated with high-risk sepsis patients, even though all patients may not be diagnosed with sepsis given the current definition and cutoffs.

**Figure 3.**
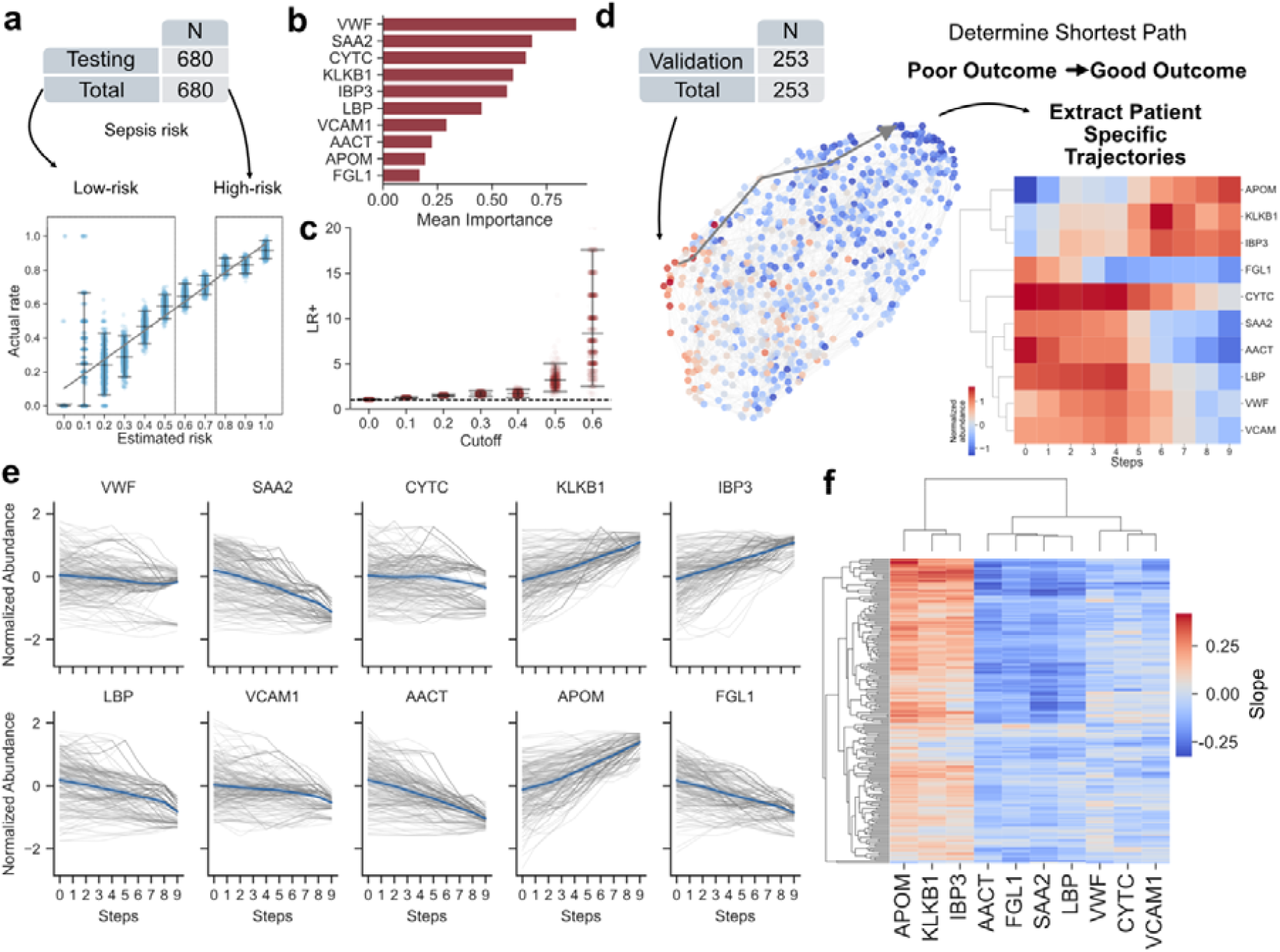
Digital twin models uncover hidden phenotypes and enable personalized treatment suggestions. **a)** Patients in the testing cohort were split into low-risk (≤0.5) and high-risk (≥0.80) groups depending on their estimated rate of developing sepsis using the digital family model from **Figure 2c**. **b)** The mean importance of the top-10 most important proteins associated with predicting high-risk sepsis. The proteins are named as follows: VWF - von Willebrand factor, SAA2 - Serum amyloid A-2 protein, CYTC - Cystatin-C, KLKB1 - Plasma kallikrein, IBP3 - Insulin-like growth factor-binding protein 3, LBP - Lipopolysaccharide-binding protein, VCAM1 - Vascular cell adhesion protein 1, AACT - Alpha-1-antichymotrypsin, APOM - Apolipoprotein M, FGL1 - Fibrinogen-like protein 1 **c)** Positive likelihood ratios (LR+) at increasing risk levels calculated by the digital twin model for the validation cohort and 95% prediction intervals. The cutoffs were determined using the median risk scores with increases based on the incremental median absolute deviation. **d)** A selected representation of an extracted pathway of a new patient from the validation cohort through the digital twin model from an area of poor outcome, defined by persistent organ dysfunction, to an area of good outcome, defined by a region where all surrounding neighbors have low (< persistent organ dysfunction mean). A molecular profile, based on the 10 proteins in the model, for the patient is extracted to provide a personalized trajectory for the high-risk patient. **e)** Line plots of the extracted protein trajectories for the 10 proteins used to build the model for each patient in the validation. The gray lines represent the trajectories of each individual patient, and the blue lines represent the average trajectory of the protein. **f)** The trends (slopes) of the proteins for each patient clustered and displayed in a cluster map. The clustering is determined by the calculated slope for all extracted at-risk patients in the validation cohort. The columns represent proteins, and the rows represent patient samples.

These high-risk non-sepsis patients, who have similar prognostic outcomes to high-risk sepsis patients (**Figure 2**), would have been difficult to identify outside the digital twin framework presented here. From the measured plasma proteome, we then employed feature selection using explainable machine learning to select a panel of the top 10 proteins that are predictive for high-risk sepsis (**Figure 3b**). Some of the selected proteins, such as Lipopolysaccharide-binding protein (LBP), and von Willebrand factor (VWF) have already been associated with severe sepsis in previous studies^42–46^, providing orthogonal evidence towards the validity of our feature selection methods. We then tested whether these selected proteins could elucidate a clustering structure in the data using k-means clustering. However, similarly to the analysis performed on the clinical data in **Extended Data Figure 1**, there was also no clear and obvious clustering structure found in the plasma proteome data (**Extended Data Figure 3f-g**). These findings provide additional evidence that digital twin modelling is a promising orthogonal approach to classic subphenotyping to identify molecularly similar patients at the time-of-admission.

### DT models can enable personalized treatment suggestions

As we observed that patients with similar clinical features were adjacent within the network (**Figure 1 and Extended Figure 2**), we investigated the possibility of tracing molecular trajectories through the digital twin model to suggest therapeutically actionable pathways. To test this, we used the panel of 10 high-risk sepsis proteins and the testing cohort (n=680) (**Figure 3b**) to build a sepsis molecular digital twin model (MDTM) (**Figure 3d**). Introspection of the network structure showed that patients with similar diagnostic (sepsis and infection) and prognostic outcomes (Persistent organ dysfunction, 30-day mortality, and ICU admission) aggregate together (**Extended Data Figure 4a**).

For each new patient in the validation cohort (n=253), we determined the prognostic risk of persistent organ dysfunction, (SOFA score >= 2 both on day 2 and day 3) and found that patients with higher risk could be more than 10 times more likely to develop persistent organ dysfunction (**Figure 3c**). For the high-risk patients, we calculated the shortest path through the digital twin model to move a patient from an area of high-risk persistent organ dysfunction to an area of low-risk persistent organ dysfunction and extracted the individual patient trajectories for each of the 10 proteins in the model (**Figure 3d**). Although there are general trends associated with some of the protein trajectories, the magnitude and direction of change were individual (**Figure 3e**), and the abundances of the 10 proteins were distributed differently throughout the MDTM (**Extended Data Figure 4b**). In general, most proteins were broadly associated with persistent organ dysfunction between individual patients, whereas proteins such as von Willebrand factor (VWF), Cystatin-C (CYTC), and vascular cell adhesion protein 1 (VCAM1) displayed more variable responses, contributing to individualized trajectories (**Figure 3f**). These proteins could act as direct targets for treatment or as proxy proteins that implicate their associated biological pathways as targets. For example, in our model, lowering VWF and FGL1 levels are linked to better predicted outcomes. Excess VWF with reduced ADAMTS13 activity promotes thrombosis and multi-organ dysfunction^44^, suggesting that ADAMTS13-based^47^ or VWF-inhibiting therapies may benefit high-VWF patients. Similarly, in our model, lower FGL1, an immune checkpoint ligand for LAG-3 that drives T-cell dysfunction^48^, predicts better patient outcome. T-cell dysfunction is a well-described pathophysiological mechanism in sepsis^49^, highlighting FGL1 as a potential therapeutic target for certain patients in sepsis. These findings demonstrate that molecular digital twin models can identify potential therapeutically actionable pathways associated with persistent organ damage for certain individual patients at the time-of-admission.

### DT models improve with additional data

To verify that digital twin models can generalize to new unseen data, we analyzed an external cohort (n=350) comprised of patients that do not have the same inclusion criteria as the sepsis alert cohorts (**Figure 1d**) and evaluated the ability of the MDTM to provide diagnostic and prognostic utility to data outside of the original distribution. Using the same 10 proteins described above, we built a MDTM using the testing cohort (n=680) and validated the diagnostic and prognostic capabilities of the digital twin model on the external cohort (**Figure 4a**). Visualization of the projected network displays distinct community separation of sepsis, infection and 30-day mortality risk (**Figure 4b**). In the external cohort, the MDTM calculated rates of sepsis, infection and 30-day mortality were statistically correlated with increased likelihood of each outcome (**Figure 4c**). These results demonstrate how digital twin models can adapt and generalize to new data with different pathological landscapes not explicitly included in the underlying data used to build the model.

**Figure 4.**
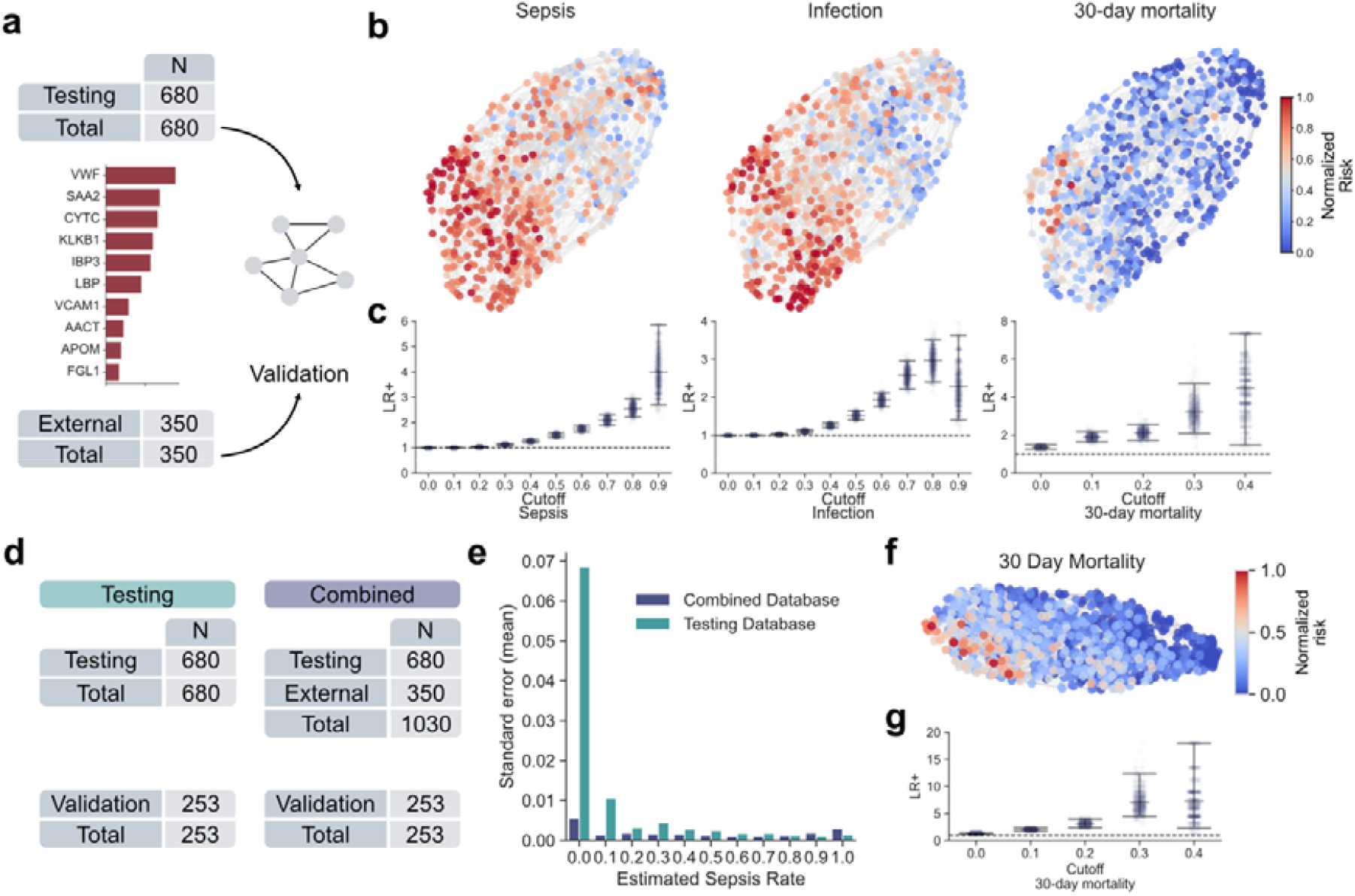
Digital twin models adapt and improve with the addition of new data. **a)** An overview of the framework used to test the generalizability of digital twin models to new data. A model was built using the 10 most important high-risk sepsis proteins from **Figure 2b** with the testing cohort (n=680), and the external cohort (n=253) was used for validation. **b)** Network projections of the underlying nearest neighbor graph of the digital twin model using the testing data and colored by the local frequency calculated using the 10 nearest neighbors in the graph. This shows an aggregation of sepsis, infection, and 30-day mortality rates aggregated in certain areas of the map. The nodes are colored based on a normalized frequency (0.0-1.0) calculated using 1000 bootstrap iterations of their 10 nearest neighbors. **c)** Positive likelihood ratios at increasing risk levels for the diagnostic and prognostic models in the external cohort and 95% prediction intervals. The dotted line is at a value of 1, where likelihood ratios above this indicate an elevated risk of association with the outcome. The risk score cutoffs are determined using increase increments of 0.1 from a baseline of 0.0. **d)** Overview of digital twin models used to test if the inclusion of the external cohort improves certain predictions. One was built using the testing corhot, and the other was built using a combination of the testing and external cohorts. The validation cohort was used for evaluation. **e)** Bar plot of the standard error of the mean values at each estimated sepsis rate for the testing and combined databases from **d**. **f)** A projected map of the digital twin model with the combined database, colored by regions of 30-day mortality. **g)** Likelihood ratios at increasing risk levels of 30-day mortality in the validation cohort and 95% prediction intervals.

As it is well known that subphenotype definitions may change when additional data is considered, we also hypothesized that digital twin models can improve their predictions as more data from different pathological distributions are included in the patient database. To test this, we evaluated if the inclusion of the external cohort in the patient database would improve the diagnosis of sepsis rates in the validation cohort. Using the same molecular digital twin architecture described above, we built two digital twin models, one using the testing cohort (n=680) as a patient database and the other a combination of the testing and external cohorts (n=1030) as the patient database and evaluated both models on the validation cohort (n=253) (**Figure 4d**). The inclusion of the external cohort, which has proportionally less patients diagnosed with sepsis, decreased the standard error of the mean overall, particularly in the lower risk value range (0.0-0.7), indicating that the inclusion of more non-sepsis patients improved the diagnostic accuracy of the model (**Figure 4e**). This model provides distinct separation between high and low-risk 30-day mortality (**Figure 4f**) with higher average likelihood of mortality at high-risk levels (0.3 & 0.4) (**Figure 4g**) compared to the base model (**Figure 4c**). These results indicate that the digital twin model can adapt and scale with new data to improve predictive performance.

### DT models may determine infection site and host pathogen response

The flexible nature of digital twin models allows the simultaneous integration of clinical and proteomics data, which can be leveraged to obtain clinically actionable insights. In sepsis, a critical step toward effective patient care is the identification of the infection site and invading pathogen, as this may directly influence antibiotic selection. As this step can take time via microbiological cultures in the clinic, prediction of infection loci and pathogen type from the host proteome at the time-of-admission could provide substantial clinical value for suggesting early antibiotic use in certain groups of patients.

To incorporate clinical outcomes with maximal proteome measurements, we performed supervised neighborhood component analysis (NCA) with a combination of the training and testing cohorts (n=1360) to project the full proteome down to a single value for 8 different clinical outcomes associated with pathogen type and infection loci (**Figure 5a**). To mitigate issues with statistical power, we focused on broad categories, such as infection loci and gram positive or negative status, that could be used to infer pathogen type and still be associated with distinct antibiotic therapies. Using these 8 infection components, we built an infection digital twin model (IDTM) and evaluated the diagnostic and prognostic capabilities of this model using the validation cohort (n=253) (**Figure 1b**). A network projection of the IDTM shows that the IDTM aggregates patients with similar pathogen type together (**Figure 5b**) and could predict the presence of a gram-positive or gram-negative infection already at the time-of-admission, with a sensitivity of 0.83 and 0.95 for gram-positive and gram-negative infections, respectively, in the combined validation cohort. Additionally, increased gram-positive or gram-negative risk scores were statistically associated with an increased positive likelihood of the respective infection, indicating increased confidence and precision at higher risk levels (**Figure 5c**).

**Figure 5.**
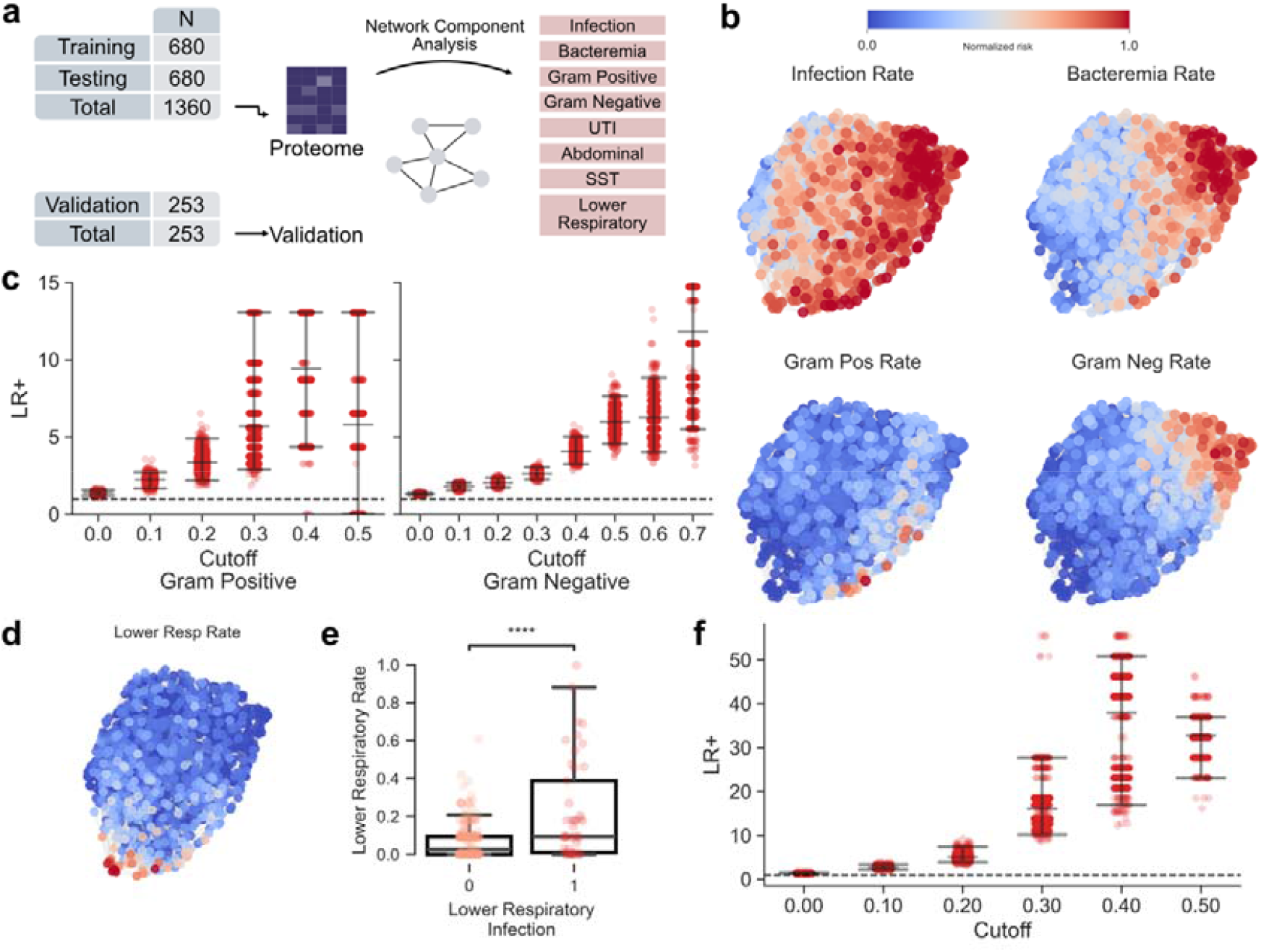
Infection location and invading pathogen determination. **a)** Overview of the data used to evaluate infection location and pathogen type determination. A combination of the training and testing cohorts were used to build the digital twin model, using a supervised projection of the proteome into 7 components related to infection. The validation cohort was used for evaluation **b)** Projected networks of the infection digital twin model. The color scale represents the normalized frequency of infection, bacteremia, gram-positive, or gram-negative for each node based on a bootstrapped estimation of the surrounding 10 nodes. **c)** Positive likelihood ratios at increasing risk thresholds for gram-positive and gram-negative infection predictions using the digital twin model and 95% prediction intervals. The risk score cutoffs are determined using increase increments of 0.1 from a baseline of 0.0. **d)** Project network infection digital twin model cohort colored by the local rate of lower respiratory infection. **e)** Lower respiratory risk values for patients with determined lower respiratory infections. The **** indicates a p-value <= 1.00 x 10^-4^ for an independent samples t-test (p-value = 3.110 x 10^-10^). **f)** Positive likelihood ratios at increasing lower respiratory infection risk values calculated by the digital twin model.

For certain types of infections, detecting the pathogen is difficult, and diagnosis relies on clinical examinations and diagnostic imaging. The IDTM also enabled the identification of many patients with lower respiratory infections that are not detected in blood culture (**Figure 5d**), with a sensitivity of 0.78 in the validation cohort. The calculated lower respiratory infection risk was statistically higher in patients with lower respiratory infection in the validation cohort (**Figure 5e**), and increased risk scores were statistically associated with higher positive likelihood ratios for lower respiratory infection (**Figure 5f**). Although there is room to optimize the sensitivity, this represents a proof-of-concept application of digital twin models to detect infections that may not be present in the blood using the host-response in the plasma proteome of a patient on admission to the emergency department.

Overall, these results demonstrate that digital twin models in sepsis may potentially be used to determine the infection site and pathogen type from the host plasma proteome already on admission to the emergency department, thereby identifying patients that could possibly benefit from narrow-spectrum antibiotics.

### DT models predict personalized vasopressor administration

Along with infection, organ dysfunction is a key hallmark of sepsis, and patients may present to the emergency department with many different types and combinations of organ dysfunction. In certain cases, patients can develop circulatory shock, which has a high association with mortality, and requires intensive care and vasopressor treatment. Analogous to the prediction of infection loci and pathogen type in **Figure 5**, we postulated that a digital twin model that incorporates clinical annotations associated with different types of organ dysfunction can be used in tandem with blood proteome measurements to predict which patients will require vasopressor treatment already on admission to the emergency department, and before fluid challenge (**Figure 6a**).

**Figure 6.**
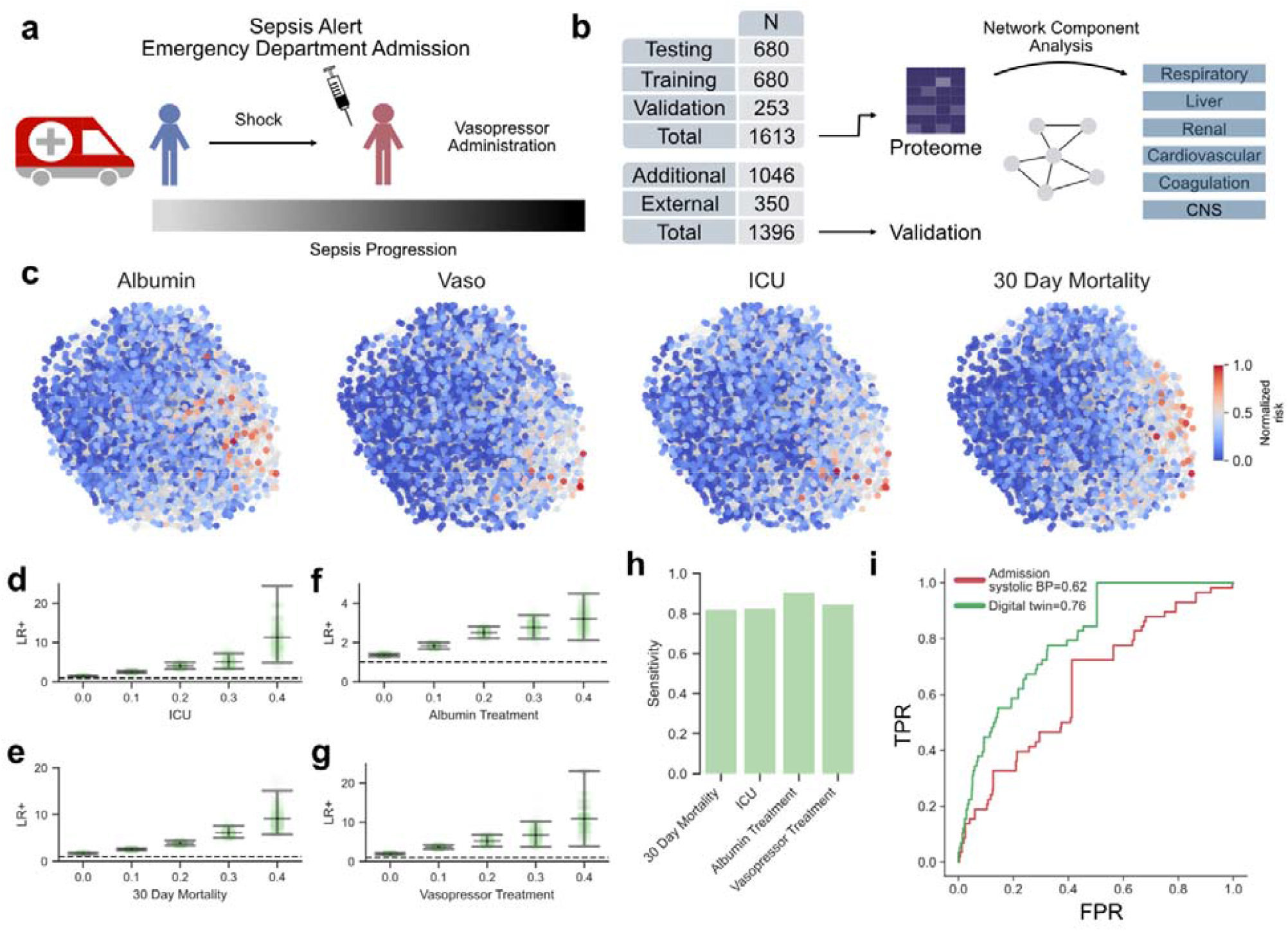
Digital twin models suggest personalized vasopressor treatment options. **a)** A visual representation of the clinical scenario for predicting patients who require vasopressor treatment. Patients suspected of sepsis who develop circulatory shock are typically transferred to the ICU and receive vasopressor treatment. **b)** Overview of the data used to evaluate the organ dysfunction digital twin model that is used to predict vasopressor treatment. The training, testing, and validation cohorts (n=1613) are used to build the organ dysfunction digital twin model using organ dysfunction-informed supervised projections of the proteome. The external and additional cohorts (n=1396) are used for evaluation of the model. **c)** Network projects of the digital twin model colored by the normalized frequency of albumin treatment, vasopressor treatment (Vaso), intensive care unit (ICU) admission, and 30-day mortality. **d)** Positive likelihood ratios for increasing risk scores of ICU admission with 95% prediction intervals. **e)** Positive likelihood ratios for increasing risk scores of 30-day mortality with 95% prediction intervals. **f)** Positive likelihood ratios for increasing risk scores of albumin treatment with 95% prediction intervals. **g)** Positive likelihood ratios for increasing risk scores of vasopressor treatment with 95% prediction intervals. **h)** Sensitivity of the digital twin model to predict 30-day mortality, ICU admission, albumin treatment, and vasopressor treatment in the combined validation set depicted in **a**. **i)** ROC curve comparing the prediction of future vasopressor treatment using systolic blood pressure at the time-of-admission to the digital twin model at the time-of-admission. The AUC for systolic blood pressure = 0.615 and for the digital twin model = 0.762. False positive rate (FPR) is plotted against true positive rate (TPR)

To test this, we sought to incorporate the proteomic profiles associated with six different organ specific SOFA scores into a single model (**Figure 6b**). Similar to the IDTM, the model was built using the 6 NCA components to determine supervised organ dysfunction components using the training, testing, and validation cohorts (n=1613). The external and additional cohorts were used for evaluation (n=1396) (**Figure 6b**). In this model, patients with different types of organ dysfunction were located in specific spatial areas of the network (**Extended Data Figure 5a**). These findings demonstrate that the blood plasma proteome has distinct protein profiles indicative of specific organ dysfunction, which can be determined based on weights extracted from the NCA (**Extended Data Figure 5b**) and validated by previous associations with a given organ system. For example, von Willebrand factor (VWF), a blood glycoprotein that promotes primary hemostasis, was important in cardiovascular dysfunction. Pulmonary surfactant-associated protein B (PSPB), a pulmonary surfactant protein highly enriched in the lung^50^, was the most important protein in respiratory dysfunction, and Cystatin C (CYTC), a well-established biomarker for chronic kidney disease, was found to be the most important protein in renal dysfunction.

The organ dysfunction-informed DTM was also able to aggregate patients together with similar 30-day mortality and ICU admission, as well as group together patients who received albumin and vasopressor treatment (**Figure 6c**). Model evaluation showed that the prognostic positive likelihood ratios for ICU admission and 30-day mortality increase substantially with risk score (**Figure 6d-e**) together with the predictive treatment scores for albumin and vasopressor treatment (**Figure 6f-g**). According to this data, patients with a vasopressor risk score of 0.4 are up to 10-times more likely to receive vasopressor treatment, suggesting a mechanism to accurately identify new patients in need of vasopressors already at the time-of-admission to the emergency department. Additionally, the organ dysfunction-informed DTM could predict 30-day mortality, ICU admission, albumin treatment, and vasopressor treatment with a sensitivity of 0.82, 0.82, 0.90, and 0.84 respectively in the combined evaluation set (n=1613) (**Figure 6h**). Compared to clinical parameters at the time-of-admission to the emergency department, such as systolic blood pressure, the vasopressor risk calculated by the model was a stronger predictor of future vasopressor treatment (24% higher ROC-AUC score than systolic blood pressure) (**Figure 6i**). Overall, the organ dysfunction-informed DTM could identify patients who received vasopressor treatment, while uncovering other clinically and molecularly similar patients who may benefit from a similar therapy. These findings highlight that time-of-admission organ-dysfunction proteome profiles can help identify informative patterns that uncover high-risk patients in need of ICU care and vasopressor treatment. This general approach could be applied towards other treatment types, potentially unlocking the potential for precision medicine in sepsis.

## Discussion

In complex and heterogeneous syndromes, such as sepsis, digital twin modelling provides an interpretable framework for patient stratification and prognostication on admission to the emergency department. As sepsis is difficult to detect outside of the ICU before repeated measurements are available, these models provide diagnostic and prognostic potential and can theoretically identify personalized therapeutic pathways for new patients. Digital twin models can embrace the heterogeneity of sepsis and inherently account for molecular and clinical diversity among patients with similar outcomes. Unlike rule-based approaches, the model can identify intricacies in the data that may not be accounted for when using subphenotypes to stratify patients suspected of sepsis. A potential extension of this framework would be to combine digital twin modelling with existing subphenotyping strategies to account for the gradients within the defined subphenotypes. Patients could first be assigned a broad subphenotype, or endotype, and then integrated into a subphenotype-specific digital twin model for fine-tuned predictions.

As sepsis is full of rare events, it is crucial that the data used to build the model accurately represents a complete picture of the pathological landscape. At the core of this study, the population scale of our cohort allows the underlying database to more closely resemble the actual population of potential patients. This is particularly important to consider before such a model would be put into use in a hospital to drive treatment. Theoretically, the clinical digital twin model (CDTM) could be immediately applied in a hospital to identify high-risk patients early and institute preventative measures, as only common and accessible parameters are used to build the model. The prognostic risk values calculated by the CDTM could be directly actionable in the clinic as they indicate patients with high 30-day mortality and ICU admission. As patient prognosis is dependent on early recognition and treatment, these results implicate the instant practical potential of digital twin models in sepsis.

A key strength of the digital twin model is the ability to integrate multiple clinical outcomes to provide actionable prognostic predictions. For example, when a patient has a high predicted risk of adverse outcomes, such as mortality or ICU admission, the cost of an incorrect treatment decision increases, as the patient is most likely severely ill. In such situations, initiating broad antimicrobial therapy and escalating care, including admission to a higher level of monitoring, could be recommended. Conversely, in clinically stable patients with a high predicted likelihood of a specific underlying infection, the model may support narrowing antimicrobial therapy and selecting a more targeted treatment strategy.

As we have shown in our results, as more data becomes available, this model could grow and adapt the underlying database to further improve prognostic accuracy and clinical utility, including the instant accommodation of new variations of sepsis when emerging diseases, such as COVID-19 or invasive group A streptococcal disease, surge. However, as this is an observational study, the direct clinical impact of digital twin models needs further prospective evaluation. Few computational tools for sepsis care are prospectively validated^51–54^, so this represents an important next step before clinical implementation.

Although all data in this study is technically available at the time-of-admission, there are some obstacles that remain before mass spectrometry-based proteomics can be applied in a clinical setting. In general, mass spectrometers require expert knowledge, can be expensive, and require substantial resources and infrastructure to operate, all which hinder the widespread adoption for routine clinical practice. Further research is also required to determine which type of model configuration is the most clinically relevant. As we have shown, different model configurations can be used to address specific questions, but the potential of a universal model has not yet been explored. Further, the prediction of pathogen type and infection location would improve significantly if the depth of the analyzed plasma proteome was improved to better reflect the host response. In an ideal scenario, and once proteomic measurements can be acquired in routine clinical practice the practical application of digital twin models could be numerous.

As an extension, digital twin models could also be used in adaptive clinical trials to select homogeneous patient groups and monitor treatment response over the course of the trial through repeated sampling. This is particularly relevant for sepsis, where over 100 clinical intervention trials have failed in the past four decades^2^. Potentially, some of these treatments could be rescued and applied to patients who present a matching receptive profile. This model could effectively be used to identify treatable traits for failed sepsis therapies in existing randomized control trials (RCTs) and test them prospectively. This future application is implied in our results, where we demonstrate that we can theoretically suggest individualized therapeutic pathways for patients, predict who may benefit from narrow-spectrum antibiotics, and identify those who will require vasopressor treatment and intensive care. These predictions are already possible on admission to the emergency department, exposing a potential mechanism for true personalized medicine in sepsis.

Overall, we provide a robust and interpretable framework for the dynamic stratification of patients suspected of sepsis at the time of admission to the hospital using adaptive digital twin models. In addition to being highly effective for the stratification of patients suspected of sepsis, we believe this methodology could be applied to other complex diseases to aid in translational research and precision medicine.

## Methods

The standards for reporting of diagnostic accuracy (STARD) guidelines were followed^55^ .

### Sepsis alert cohort settings and patient population

Patients were eligible for inclusion if ≥ 18 years and admitted to the emergency department (ED) at Skåne University Hospital, a tertiary hospital in Lund, Sweden as sepsis alert. Sepsis alert is a modified triage system to detect patients with suspected sepsis in the ED and to provide instant care^56^ . These types of triage systems are becoming more common although with different inclusion criteria. The criteria for sepsis alert at Skåne University Hospital are patients with any of the following: active seizures, unconsciousness, a respiratory rate higher than 30 breaths/min or lower than 8 breaths/min, oxygen saturation below 90%, a regular heart rate over 130 beats/min, or an irregular heart rate over 150 beats/min, a systolic blood pressure (SBP) below 90 mmHg, p-lactate >3.5 mmol in combination with fever, >38°C or a history of fever or chills^57^. Patients meeting the criteria for sepsis alert are cared for immediately by emergency physician, infection consultant, nurse, and assistant nurse and undergo immediate control of vital parameters, physical examination, blood sampling for biochemical analyses and culture and other microbiological sampling. Sepsis alert patients are also considered for other diagnostic procedures, treatment, level of care and surveillance.

Patients were included prospectively, consecutively from 1st September 2016 to 31st of March 2023 and venous blood samples were drawn in citrate tubes at admission. Samples were centrifuged and stored at -70°C within 2 hours of collection.

### External cohort settings and patient population

Patients considered for inclusion were part of a mi, multicenter, observational study using a convenience sample of ED patients previously reported^58^. Only those patients enrolled at Skåne University Hospital were included in this study. Recruitment occurred in February 2015 and then again from January to March 2016. Inclusion criteria for patient enrollment were as follows: age > 18 years and at least one of the following conditions irrespective of the underlying cause: respiratory rate > 25 breaths per minute, heart rate > 120 beats per minute, altered mental status, systolic blood pressure < 100 mm Hg, oxygen saturation < 90% without oxygen, oxygen saturation < 93% with oxygen, or reported oxygen saturation < 90%.

Blood samples were collected in EDTA (Ethylenediaminetetraacetic acid) tubes at the time of enrollment in the emergency department and were centrifuged and stored at –80°C within 2 hours of collection.

### Ethical permission

For the sepsis alert study cohorts (testing, training, and validation cohorts), informed consents were collected through an opt-out procedure. Patients received a mail following the ED admission and were given the opportunity to opt-out of inclusion in the study by sending back a form in a pre-franked envelope. Ethical approval for the study was obtained from the regional ethics board (file numbers 2022-01454-01, 2014/741 and 2016/271).

The ethical permits for the external cohort are file numbers 2014/741, 2017/26 and 2017/106.

### Sepsis Alert clinical data collection

For the training, testing and validation cohorts clinical data was collected retrospectively through a manual and structured in-depth clinical chart review. Chart reviews and validations were conducted by clinicians working in acute care settings with sepsis patients. Prior to undertaking these assessments, all reviewers received appropriate training and adhered to a structured protocol (see **Extended Data 1** for the structured protocol and case report form (CRF) completion guidelines). None of the reviewers were aware of any additional findings (e.g., proteomic data) at the time of the chart review.

Data were collected on demographics, vital signs, laboratory tests, microbiological investigations, medical history, diagnostic and therapeutic procedures, concomitant or newly initiated medications, and level of care. The likelihood of organ dysfunction, infection, and sepsis was determined according to the Sepsis-3 criteria, with organ dysfunction defined as a ≥ 2 increase in the Sequential Organ Failure Assessment (SOFA) score from baseline. Infection was assessed primarily using the LMCI score. The SOFA score was modified for compatibility with use outside intensive care units (ICUs) (**Supplementary Table T2**). SOFA scores were calculated at baseline and every 24 hours during the first 72 hours of admission.

In the context of clinical diagnosis, sepsis is conditional on infection, and although there have been multiple efforts to create definitions for infection in sepsis, there currently is no accepted gold standard. Clinical adjudication has demonstrated poor interrater agreement, which facilitated the use of LMCI in combination with adjudication as a framework for infection in our study to minimize the inclusion of non-sepsis patients^59–61^. The different cohorts mirror different diagnostic approaches in sepsis research, such as clinical validation by criteria and adjudication, adjudication solely, or International Classification of Diseases (ICD)-diagnosis.

Based on this strategy, five groups were defined within the cohort: (1) no organ dysfunction and no infection; (2) organ dysfunction without infection; (3) no organ dysfunction with infection; (4) sepsis; and (5) septic shock, classified according to the Sepsis-3 definitions.

Medical records are available for all acute care hospitals and all but one of all hospitals in the region. For the different components of the SOFA score the baseline was defined as the latest value measured before the present admission but within a year. For patients without values measured the year prior, SOFA score was assumed to be zero in patients not known to have preexisting organ dysfunction. Preexisting organ dysfunction was primarily measured by parameters and secondarily by previous diagnoses.

The LMCI score was used as decision support for the clinical chart review but could be rejected at adjudication. In cases of uncertainty, clinical data were validated by a second physician. Validation refers to conducting a second round of chart review on patients primarily to confirm infection and organ dysfunction diagnoses. In cases of diagnostic uncertainty *i*.*e*. when the clinical chart reviewer adjudicated the infection or sepsis diagnoses differently than the LMCI or SOFA, adjudication was discussed with 1 of 2 senior physicians. The following comorbidities were recorded: cardiovascular disease, liver disease, malignancy, respiratory disease, diabetes mellitus, chronic kidney disease, immunodeficiency, and Charlson comorbidity index.

For the additional cohort, partial clinical data were extracted automatically from electronic health records. Data used in analyses which were focusing on prognostic outcome, such as intensive care and vasopressor administration were manually validated.

Microbiological diagnostics were performed at the clinical microbiology department (Laboratory Medicine Skåne, Lund, Sweden) per standard clinical practice.

### External cohort clinical data collection

Clinical data was collected retrospectively through a detailed review of clinical charts. This process was conducted by medical researchers and subsequently validated by infectious disease clinicians. Data encompassed a wide range of parameters, including demographics, vital signs, laboratory results, microbiological investigations, medical history, diagnostic and therapeutic procedures, concomitant or new medications, and level of care.

For the calculation of the Sequential Organ Failure Assessment (SOFA) score, a modified SOFA-scale was employed as described in the reference^58^ . The maximum SOFA value within the initial 72 hours was determined for each patient.

The presence of an infection was assessed independently by two infectious diseases (ID) consultants. In instances where their assessments differed, a third ID physician reviewed the data, and a consensus was reached through discussion among all three physicians. Patients were categorized into four groups: verified infection, probable infection, probably not an infection, and no infection. Patients who had a maximum SOFA score of ≥ 2 and were classified as having either a verified or probable infection were considered to have sepsis or septic shock. The classification above was used for external validation.

The diagnosis group descriptions for the external cohort (**Figure 1d**) were aligned with the five classes used for the sepsis alert study cohort patients since the external cohort was collected when the sepsis-2 definition was in use. Patients with infection and organ dysfunction receiving either epinephrine or norepinephrine within the first 72 hours were classified as having septic shock, regardless of the reason for drug administration (hypotension or other reason), blood pressure or, lactate level. Patients that died during the 72-hour period were adjudicated to respective group based on recorded values. For details see **Supplementary Table T1c**.

### Clinical data preprocessing

The sepsis alert study cohort consisting of 1360 samples was split into 2 groups (50%-50%) both consisting of 680 patients. The split was stratified to preserve the distributions of sepsis diagnosis groups in the original 1360 patients. These groups became the training and testing cohorts. The validation cohort and the external cohort were not filtered further.

The 13 time-of-admission clinical parameters used throughout the study are: SaO_2_, respiratory frequency, heart rate, systolic blood pressure, mean arterial pressure, mental status, temperature, creatinine, bilirubin, c-reactive protein, platelets, sex, and age. Missing values for the continuous clinical parameters SaO_2_, respiratory frequency, heart rate, systolic blood pressure, mean arterial pressure, temperature, creatinine, bilirubin, c-reactive protein, platelets, and age were imputed with mean values from each parameter if needed. These values were also scaled to unit variance by subtracting the mean and dividing by the standard deviation. Mental status in the ambulance was imputed with 0 if missing.

### Cluster analysis

Cluster analysis was performed using the 13 time-of-admission clinical parameters listed above. Briefly, we utilized consensus clustering, using k-means clustering^5,7,8^, to optimize the number of clusters within our training and testing cohorts. First, we performed Ordering Points To Identify the Clustering Structure (OPTICS) to investigate if the data had a clear clustering structure. As the data was not found to have a clear clustering structure, we performed partitioning of the data using consensus clustering with k-means for different numbers of clusters (k=2-11). We ran 1000 iterations at each k, subsampling the data to 90% for each iteration and performed k-means clustering. A final hierarchical clustering is then used on the results of the subsampling iterations to determine a final cluster membership for each patient. An implementation of this algorithm is available on GitHub (https://github.com/InfectionMedicineProteomics/DigitalFamilyAnalysis). We then analyzed the empirical cumulative distribution function (ECDF), the delta between the areas under the curve (AUC), and silhouette score analysis to determine the optimal number of clusters in the data. As the delta-AUC was minimal for k=4, the silhouette score was reasonably high, and the cluster passed visual inspection, k=4 was selected as the optimal number of clusters for the data.

Using the top-10 proteins for high-risk sepsis, we also analyzed the potential clustering structure for more than 300 clusters using k-means clustering. We plotted the within-cluster sum of squares (WCSS), which is an estimation of cluster quality, and plotted this for all k. Using the elbow method, we then determined the optimal number of clusters based on the WCSS values and found this to be 44. We also plotted the silhouette scores for all k.

### Sample preparation

Plasma samples were diluted 1:10 with 100 mM ammonium bicarbonate (Sigma-Aldrich), and a total of 10 μl of diluted sample (corresponding to 1 μl plasma) was used for protein digestion. Samples were incubated for 60 minutes at 37°C in 4 M urea (Sigma-Aldrich) and 60 mM dithiothreitol (DTT, Sigma-Aldrich) for denaturation and reduction. Addition of DTT to the samples, as well as the following additions, were performed with an Agilent Bravo liquid handler. The samples were then alkylated using 80 mM 2-iodoacetamide (Sigma-Aldrich) for 30 minutes at room temperature in the dark, followed by digestion with 2 μg LysC (Lysyl Endopeptidase, Mass Spectrometry Grade, Wako) for two hours at room temperature. The samples were further digested using 2 μg trypsin (sequence-grade modified porcine trypsin, Promega) for 16 hours at room temperature. The digestion was stopped with 10% trifluoroacetic acid until pH ∼2. The samples were stored at -80 °C until mass spectrometry analysis.

For peptide clean-up, digested samples were diluted 1:12 in water to generate suitable concentrations and subsequently loaded onto disposable Evotip Pure C18 trap columns (Evosep Biosystems, Odense, Denmark), which were prepared according to the manufacturer’s instructions. Briefly, the Evotips were activated in 0.1% formic acid in acetonitrile, conditioned by wetting the tips in 2-propanol and equilibrated in 0.1% formic acid. Twenty microliters of each diluted sample were transferred to tips, followed by washing with 0.1% formic acid. The Evotips were stored in 0.1% formic acid until LC-MS/MS analysis.

### Mass spectrometry data acquisition

Evosep One LC system (Evosep Biosystems) was used for separation with nanoflow reversed-phased chromatography. All samples were run with the 60 samples per day (SPD) method (gradient length of 21 min, vendor standard settings) with a flow rate of 1 μl/min. MS data was acquired using data-independent acquisition serial fragmentation (diaPASEF) method. For the training and testing cohorts, samples were analyzed with timsTOF Pro 2 ion mobility mass spectrometer (Bruker Daltonics) using diaPASEF and variable windows (**Supplementary Table T3**). For the remaining cohorts, samples were analyzed with a timsTOF HT ion mobility mass spectrometer (Bruker Daltonics) with variable windows (**Supplementary Table T4**). In all cases, a 8 cm x 150 μm Evosep column (Evosep Biosystems) packed with 1.5 μm ReproSil-Pur C18-AQ particles was used. The accumulation and ramp times were set to 100 ms. Variable isolation windows were used for the diaPASEF method with an estimated cycle time of 2.76 s. The collision energy was ramped linearly as a function of the mobility from 59 eV at 1/K0 = 1.5 Vs cm-2 to 20 eV at 1/K0 = 0.6 Vs cm-2.

### Mass spectrometry data analysis

A reviewed FASTA file was downloaded from UniProtKB ((taxonomy_id:9606) AND (reviewed:true)) and filtered for the tissue specific and plasma proteins contained in the spectral library used in Malmström et al. 2025^50^, and the spectral library was created by predicting spectra, retention time, and ion-mobility with AlphaPeptDeep^62^.

Precursor quantities were extracted for the training, testing, validation, external, and additional cohorts separately. Targeted extraction was performed using DIA-NN (2.2.0)^63^ with the spectral library above and match-between runs. Configuration details are available on GitHub (https://github.com/InfectionMedicineProteomics/DigitalFamilyAnalysis/blob/main/workflows/diann.smk).

Analysis was automated and controlled using the workflow manager snakemake (v8.20.3)^64^, with the config file and snakefile available on GitHub https://github.com/InfectionMedicineProteomics/DigitalFamilyAnalysis/tree/main/workflows).

Prior to protein quantification, the main sepsis alert cohort was split according to the training and testing cohort splits in the clinical data described above. Protein quantities for each of the 5 separate cohorts were calculated separately. Results were filtered for 1.0% false discovery rate at the precursor and library (protein) level.

### Protein quantification

For each of the 4 cohorts, the extracted precursors were first normalized at the sample level using a mean normalization method. For each *j*-th sample in the quantitative precursor ion matrix:

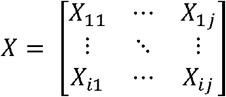

We calculated a sample-wise statistic, *S*_*j*_:

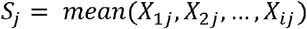

Using sample-wise statistic and the mean of the sample-wise statistic the precursor ion intensities are corrected according to the following formula:

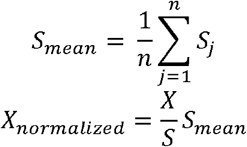

This normalization procedure was conducted using a retention-time sliding window scheme as described in NormalyzerDE^65^. The normalized signal is then *log2* transformed.

The normalized precursor ions are quantified into proteins using a modified version of the *iq* relative protein quantification algorithm^66^. Relative protein abundances are calculated from the precursor signal using only the top 5 most abundant precursors per protein. This ensures that the relative rank profiles of the quantified precursors are preserved while minimizing quantitative protein variance across samples.

Proteins were quantified from normalized precursors using a python implementation of the relative quantification algorithm *iq*^66^ using the top 5 most abundant precursors per protein. Ratios between all samples are calculated, and an over-represented system of equations is solved to quantify each protein profile per sample using linear least squares.

Proteins that were not present in a minimum of 50% of the samples per cohort were removed and missing values were imputed using a uniform distribution built from the bottom 5% of the abundance profile of the cohort.

After individual protein quantification, only shared proteins that were quantified across the 4 cohorts were kept for downstream analysis, and protein quantities were corrected to the training cohort to minimize batch effects.

Immunoglobulin proteins and heme-related proteins were removed from downstream analysis.

All algorithms are implemented in the free and open-source software package DPKS^67–69^ available on GitHub (https://github.com/InfectionMedicineProteomics/DPKS/).

### Digital twin model

The digital twin models in this study were created by first creating nearest neighbor graphs on each specific search space. The search space is partitioned into a tree data structure to allow for fast extraction of the nearest neighbors of a query sample so that all samples do not need to be considered. In this study, the 10 nearest neighbors are extracted to create each digital family for every model that is presented. This number was optimized using multiple metrics. We considered the intra-neighborhood distances and intra-neighborhood standard deviations, as well as the precision of the nearest neighbors to diagnose sepsis. Considering these metrics, we also visually inspected the structure of the resulting network to select k=10 as an optimal number of neighbors (**Extended Data Figure 1**).

Distances between the query sample and the samples in the search space are determined using the *n*-dimensional Euclidean distance, where *n* is the number of features used in the model. This distance can be expressed mathematically as follows:

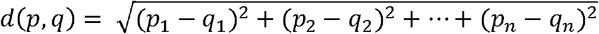

This calculates the distance between a sample *p* = (*p*_1_,*p*_2_,… *p*_*n*_) in the search space, and *q* = (*q*_1_,*q*_2_,… *q*_*n*_) the query sample. Once the nearest 10 neighbors are identified, predictions can be made by calculating probabilities for any binary clinical outcome by summing the number of occurrences of the clinical outcome in the digital family and dividing by the size of the digital family, *n*:

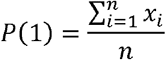

These probabilities are used to predict the rates of the clinical outcome of interest for each query patient.

To provide confidence intervals for each prediction, or prediction intervals, the background search space of the digital family model is resampled (*k* = 1000) without replacement at 90% of the search space. Predictions are used by aggregating the predictions across the sampling iterations, or bagging^70,71^ .

The source code for the digital family algorithm is available on GitHub (https://github.com/InfectionMedicineProteomics/DFModel).

### Statistical analysis

Statistical differential tests were performed between high and low risk sepsis groups using linear regression models. As the nature of the data is log2-transformed normalized protein quantities and all differential tests are between 2 groups, linear regression models were chosen. Since only 2 groups are being compared throughout this study, the assumptions of linearity between groups are upheld, all samples are independent, protein variance across samples were minimized using the relative protein quantification algorithm described above, and the distribution of residuals is assumed to be normal.

Patients in the testing cohort were split into low-risk (≤0.5) and high-risk (≥.080) sepsis probabilities based on the prediction of sepsis probability from the clinical digital twin model. We identified differentially abundant proteins between the low-risk and high-risk groups using the statistical method analysis method described above. Linear regression models were trained for each protein across samples to determine the statistical significance of the log2 fold change between low-risk and high-risk groups. As this was an exploratory analysis, and since the biological signal between groups is very strong, no optimization of the differential abundance algorithm was performed. P-values from the differential abundance analysis were corrected using the Benjamini-Hochberg false discovery rate method to perform multiple testing correction and control the false discovery rate^72^ .

Biological pathway enrichment was performed using upregulatd or downregulated statistically significant proteins (*corrected pvalue* ≥ 0.01). Overrepresentation tests using the enrichr tool^73,74^ with GSEAPY^75^ were used to find enriched biological pathways from the Gene Ontology database^76,77^. The algorithms used to perform the statistical analysis and biological pathway enrichment are available on GitHub (https://github.com/InfectionMedicineProteomics/DPKS).

Positive likelihood ratios and R^2^ values were calculated at different risk levels using the results from all 1000 bootstrap iterations. Positive likelihood ratios are defined as *sensitivity / (1 – specificity)*. Prediction intervals (from the model bootstrapping iterations) were calculated for R^2^ values and positive likelihood ratios.

### Clinical digital twin analysis

To perform clinical digital family analysis, 13 clinical parameters that are available at the time-of-admission were extracted for each patient in the training and testing cohorts. The clinical parameters used are: blood oxygen saturation (SaO_2_), respiratory rate, heart rate, systolic blood pressure, mean arterial pressure, mental status, temperature, creatinine, bilirubin, platelets, C-reactive protein (CRP), sex, and age. The training cohort was used as the search space and the testing cohort was used to evaluate the model. The clinical digital family model was used to diagnose the rate of sepsis (defined as either sepsis or septic shock) and infection (defined as a verified infection based on the LMCI score), as well as prognose persistent organ dysfunction (defined as an increase in SOFA >= 2 on day 2 or day 3), and 30-day mortality, and admission to the ICU. To evaluate the calibration of model, we binned all patients in the testing cohort based on their estimated probabilities of the 5 predictions and measured the actual rates in each bin. We calculated prediction intervals using the 1000 bootstrapped partitions of the model to calculate mean positive likelihood ratios. Sensitivity for 30-day mortality and ICU admission was evaluated using the testing cohort.

### High-risk sepsis analysis

#### Feature Selection

In high-dimensional fields, such as proteomics, too many features, or proteins, can lead to serious analytical challenges^78^. Therefore, it can be beneficial to project the proteome to a lower dimension to maximize the application of available molecular information when predicting disease or other biological outcomes^79–81^. The feature selection process used in **Figure 3** is a multi-step process with the goal of minimizing multicollinearity and selecting proteins that are the most predictive of a particular clinical outcome.

We first minimize the number of colinear proteins using maximum-Redundancy-Maximum-Relevance (mRMR)^82,83^ to first select a subset of 50 proteins. We then quantified the importance of each protein by analyzing the impact it has on the output of the model. This is done by shuffling the values for a feature across the samples in a dataset and recording how the output of the model changes. First, a baseline set of predictions, *y*, are produced, and then a feature is shuffled to produce new predictions, *ŷ*, and the mean is taken across all patients to provide a mean importance for a given protein. This can be expressed using the following equation:

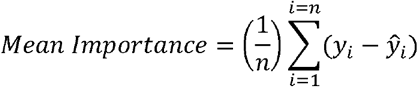

Each feature is shuffled (k=100) to provide robust estimates of protein importance. The dataset is further bootstrapped (k=100) to minimize issues caused by local variance of the background dataset. This method is similar to existing perturbation feature attribution methods and other methods such as SHAP or LIME^84,85^, but has been simplified for use with proteomics data and binary classification. The algorithm is implemented in the DPKS python package and is freely available on GitHub (https://github.com/InfectionMedicineProteomics/DPKS)^67–69^ .

The machine learning model used for feature selection is a logistic regression classification model with *l*2-regularization and balanced sample weighting to minimize issues caused by any possible class imbalances. The solver used was Large-scale Bound-constrained Optimization (L-BFGS-B) and the *l*2-regularization was controlled with *C* =1.0. The implementation used was LogisticRegression from the scikit-learn python library^86^.

The top-10 features based on importance in predicting high-risk sepsis were selected.

### Molecular Digital Twin Model

A digital twin model was built using the testing cohort and the top-10 features from the method described above. K=10 neighbors, Euclidean distance, and 1000 bootstrap iterations were selected as hyperparameters as described above. The validation cohort (n=253) was used for validation of this model and to extract patient specific trajectories.

### Patient-specific trajectories

As the digital twin model is a nearest neighbor graph, pathways between nodes can be extracted to examine how proteins change to move from one area of the graph to another. Using an implementation of the A* pathfinding algorithm^87^, we extracted the shortest pathways through the digital twin model for patients with persistent organ dysfunction in the validation cohort that were greater than the mean risk rate in the model. The heuristic for the A* algorithm is the Euclidean norm between a source node and nodes within a path for the 10 proteins (*n* =10), defined as follows:

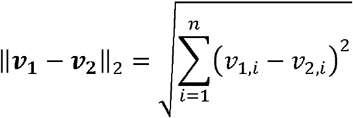

Where ***v*_1_** is the vector of 10 proteins for the source node and ***v*_2_** is the feature vector for the node within the path. Nodes in the nearest neighbor graph are traversed, with nodes that minimize the heuristic prioritized to be searched first, until a goal is reached. The goal is defined as a node where all surrounding nodes have a persistent organ dysfunction risk less than the average of the model.

### Infection location and pathogen type determination

Seven clinical outcomes were used to project the full proteome to a lower dimensional space using neighborhood component analysis (NCA)^88^ . The following comparisons were made for 7 infection type groups:

- Bacteremia: defined as a positive blood culture without the presence of irrelevant bacteremia.
- Gram positive infection: Defined as a positive blood culture for one of the following pathogens: *Staphylococcus aureus, Streptococcus pneumoniae, Beta-hemolytic streptococcus (groups a, b, c, g), Enterococcus faecalis, Enterococcus faecium*
- Gram negative infection: Defined as positive blood culture for one of the following pathogens: *Klebsiella pneumoniae, Pseudomonas aeruginosa, Escherichia coli, Enterobacter cloacae*
- UTI: Patients with urinary tract infection but no abdominal, skin and soft tissue (SST), or lower respiratory infections.
- Abdominal: Patients with abdominal but no UTI, SST, or lower respiratory infection.
- SST: Patients with skin and soft tissue infection, but no UTI, abdominal, or lower respiratory infection.
- Lower Respiratory: Patients with pneumonia, but no UTI, abdominal, or SST infection.

NCA was used to project the proteome down to 7 unsupervised components that were subsequently used to create the infection digital twin model using the method described above.

### Vasopressor treatment prediction

Seven clinical outcomes were identified and analyzed to create an organ dysfunction-informed digital twin model to predict if a patient will receive vasopressor treatment. Organ dysfunction groups based on SOFA scores (respiratory, renal, liver, cardiovascular, coagulation, and central nervous system) were selected. Patients were labeled with organ dysfunction if the increase in SOFA score based on the following increases from baseline (day 0) on days 1, 2, or 3:

- Respiratory organ dysfunction: 2
- Coagulation organ dysfunction: 1
- Liver organ dysfunction: 1
- Cardiovascular organ dysfunction: 1
- Central nervous system (CNS) organ dysfunction: 1
- Renal organ dysfunction: 1

For respiratory dysfunction the zone of uncertainty between presence and absence of organ dysfunction is expanded since oxygen saturation is a volatile examination. NCA was used to project the full proteome to 6 supervised dimensions.

Albumin treatment was indicated if the patient received any albumin treatment within the first 72 hours.

The organ-dysfunction informed digital twin model was used to predict if patients received albumin treatment, vasopressor treatment, 30-day mortality, and ICU admission. Statistical analysis and likelihood ratio determination were determined as described above.

### Predicted clinical outcome latent space

Using the NCA models for any *M* set of clinical outcomes, a latent space of size *M* is created. For a set of models:

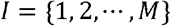

For each *i* ∈ *I*, the full proteome is projected using a linear model to single value that maximizes the classification accuracy of a nearest neighbor algorithm for the given clinical outcome.

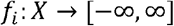

For a specific patient, *x* ∈ *X, f*_*i*_ (*x*), gives the value that the patient has a particular clinical outcome. From there, the projected clinically-informed values for all classifiers *f*_*i*_ ∈ *I*, can be stacked to form a latent vector for a patient:

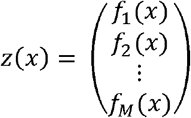

Since we know exactly what each *f*_*i*_ (*x*) is associated with, we can therefor interpret the latent vector qualitatively, as it is possible to determine which clinical outcomes a patients is predicted to have.

Additionally, we can trace back each value to a set of proteins based on the weights of the NCA.

Finally, we then stack all patient probability vectors into a dense matrix *Z* with *n* rows, for the number of patients, and *M* columns, for the number of classifiers, that can be used to build digital twin models:

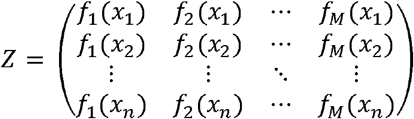

This matrix can be adapted based on which clinical outcomes are desired to connect to the proteome to build a clinically-informed digital family model.

## Data Availability

Data produced in the present study are available upon reasonable request to the authors

## Inclusions and Ethics

The ethical permits for the external cohort are file numbers 2014/741, 2017/26 and 2017/106.

## Data Availability

All mass spectrometry files and analysis results have been deposited to the ProteomeXchange Consortium (identifier PXD051271) via MassIVE (identifier MSV000094486). Clinical data and relevant patient annotations can be requested. Quantified proteins and the relevant anonymized clinical parameters to perform all analysis in the manuscript are available on GitHub (https://github.com/InfectionMedicineProteomics/DigitalFamilyAnalysis). The validation, external, and additional mass spectrometry files will be available on request. Results files are available on GitHub (https://github.com/InfectionMedicineProteomics/DigitalFamilyAnalysis).

## Code Availability

Source code related to the digital family algorithm is provided as a python package, and is available on GitHub (https://github.com/InfectionMedicineProteomics/DFModel). The code to create the figures and analysis is available on GitHub (https://github.com/InfectionMedicineProteomics/DigitalFamilyAnalysis). All code related to statistical analysis and explainable machine learning is open-source and freely available on GitHub (https://github.com/InfectionMedicineProteomics/DPKS). The workflow for running DIA-NN is available on GitHub (https://github.com/InfectionMedicineProteomics/DigitalFamilyAnalysis/tree/main/workflows).

## Acknowledgements

A special thanks to Erik Hartman for the always insightful computational discussions and to Caroline and Paul Sverdrup for their generous support. Lars M. is funded by the Swedish Research Council (grant number VR-2020-02419), the Wallenberg foundation (grant number 2016.0023) and Alfred Österlunds Foundation. J.M. is a Wallenberg academy fellow (KAW 2017.0271) and is also funded by the Swedish Research Council (Vetenskapsrådet, VR) (2019-01646 and 2018-05795), the Wallenberg foundation (KAW2016.0023, KAW2019.0353 and KAW2020.0299), Vinnova DNR 2023-04243, and Alfred Österlunds Foundation. E.M. is funded by Wenner-Gren Foundation (FT2020-0003), the Crafoord Foundation, and the Swedish Society of Medicine (SLS-985287). F.K. is funded by Region Skåne ALF project and the Crafoord Foundation. A.L. is funded by the Swedish Research Council VR 2023-02707 and Region Skane ALF project 2022-0146. Lisa M. is funded by Governmental funding of clinical research within the Swedish National Health Service (ALF) 2022:YF0025, the Swedish Society of Medicine and the Clas Goschinsky Foundation.

## Supplementary Figures

**Extended Data Figure 1.**
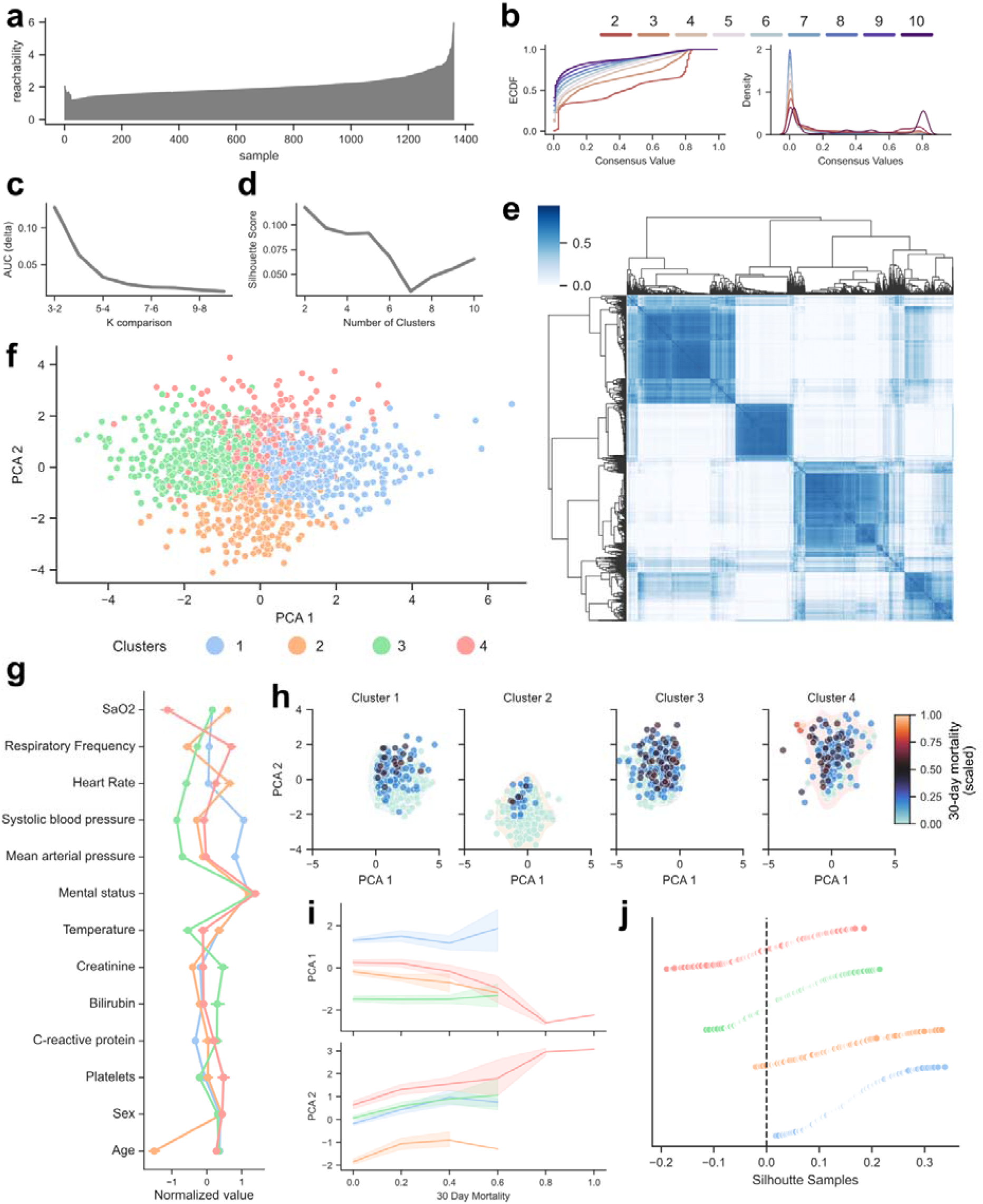
Clinically distinct subphenotypes. **a)** Ordering Points To Identify the Clustering Structure (OPTICS) analysis of the training and testing cohort combined. The results indicate no clear clustering structure. **b)** Empirical cumulative density function (ECDF) plot of different k-clusters using the consensus clustering algorithm with k-means clustering. **c)** Differences in the area under the curve (AUC) from **c** between different values of *k*. **d)** The average silhouette score for different numbers of *k*. **e)** Clustermap of the consensus clustering values using the selected k=4 clusters. **f)** The PCA plot from **a** colored by the optimally determined 4 partitions. **g)** A point plot displaying the variation of clinical parameters throughout each of the 4 subphenotypes. **h)** Projected maps of the 30-day mortality gradients within each defined subphenotype. Each node is colored by the frequency of 30-day mortality calculated by the nearest 10 neighbors in the subphenotype. **i)** Distribution of principal components on 30-day mortality. **j)** Silhoutte scores for each patient sample in each defined subphenotype.

**Extended Data Figure 2.**
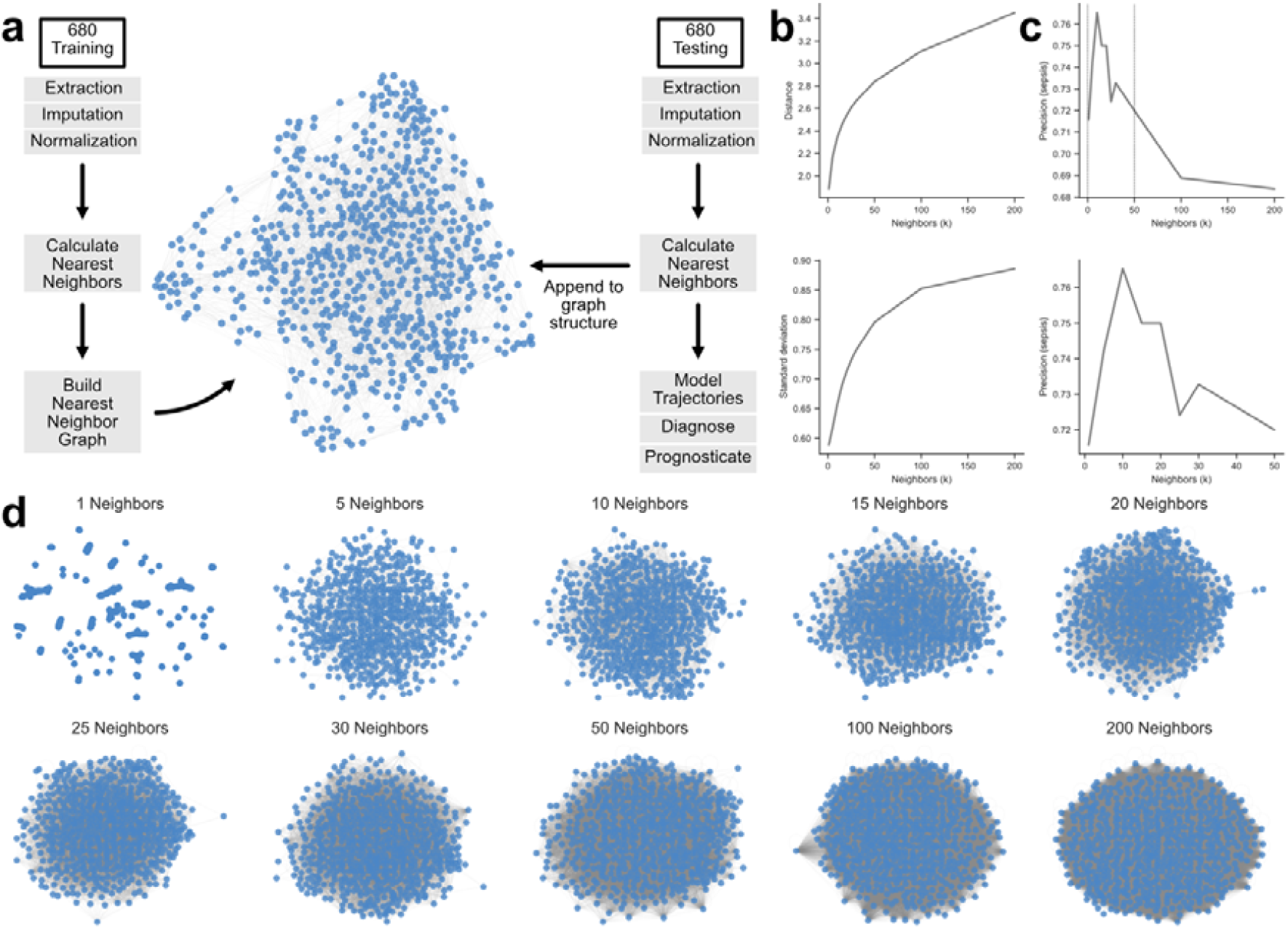
Digital twin model selection and workflow. **a)** Overall workflow for building and evaluation of the clinical digital twin model. Data is extracted, imputed, and normalized, followed by the construction of a nearest neighbor graph. This graph represents the structure of the digital twin model. For new patients, data is extracted and transformed, and the nearest neighbors are calculated for the new patient in the digital twin model. This patient is then integrated into the model, where the 10 nearest neighbors can be used to diagnose, prognose, and make predictions about future treatment. This workflow can be generalized to all digital twin models described throughout the study. **b)** Lineplots showing the relationship of intra-family distance and variation at different k-neighbors. **c)** Lineplots showing the relationship of diagnostic precision for sepsis at different k-neighbors. **d)** A projected network of the digital twin model created using different k-neighbors.

**Extended Data Figure 3.**
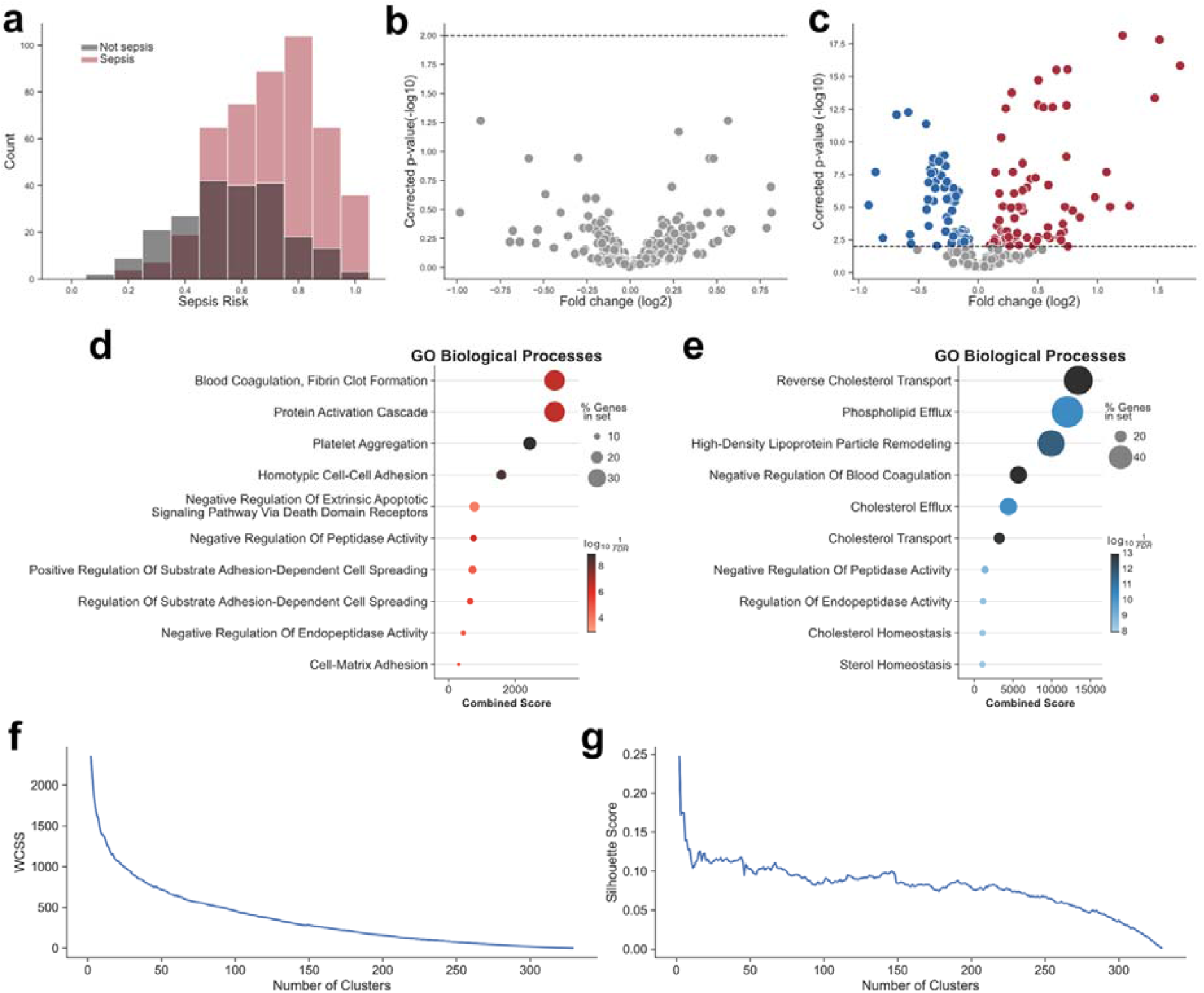
Proteomic analysis of high-risk sepsis. **a)** Histogram of the sepsis risk values calculated by the clinical digital twin model from **Figure 1**. Colored by sepsis diagnosis. **b)** Volcano plot of non-sepsis to sepsis patients within the high-risk sepsis group (>0.8). **c)** Volcano plot of low-risk compared to high-risk sepsis patients in the testing cohort. Each dot represents a protein. Red proteins are positively enriched, and blue proteins are negatively enriched. Gray proteins were not found to be statistically significant (Corrected p-value > 0.01). **d)** Gene ontology enrichment analysis of the positively enriched proteins from **c**. Dot size is determined by the percentage of proteins found in the set for the term. And the color is the log transformation of the inverse p-value. **e)** Gene ontology enrichment analysis for the negatively enriched proteins from **c**. **f)** Within cluster similar score (WCSS) across different values of k-clusters. The WCSS is used to perform elbow analysis to determine the optimal number of clusters using k-means clustering with the selected panel of 10 proteins from **Figure 3b**. **g)** Silhoutte scores at different k for optimal clustering determination using the selected panel of 10 proteins from **Figure 3b**.

**Extended Data Figure 4.**
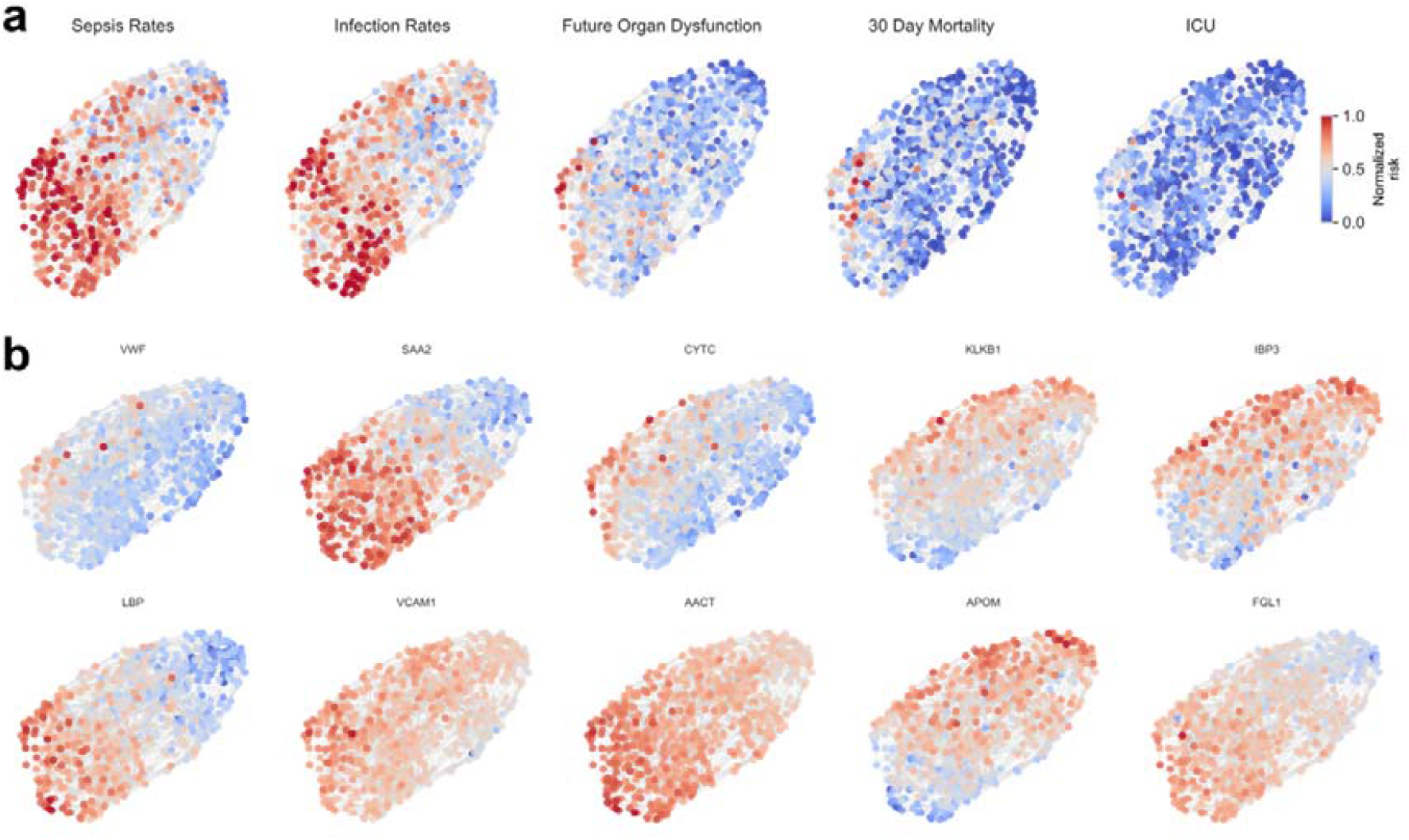
Digital twin network maps in high-risk sepsis. **a)** Projections of the high-risk sepsis molecular digital twin model colored by the local sepsis, infection, persistent organ dysfunction, 30-day mortality, and ICU admission rates. This model is built using the testing cohort and is the same model as presented in **Figure 3**. **b)** The same projections colored by the normalized mean abundances of each protein used to build the molecular digital twin model from **Figure 3**.

**Extended Data Figure 5.**
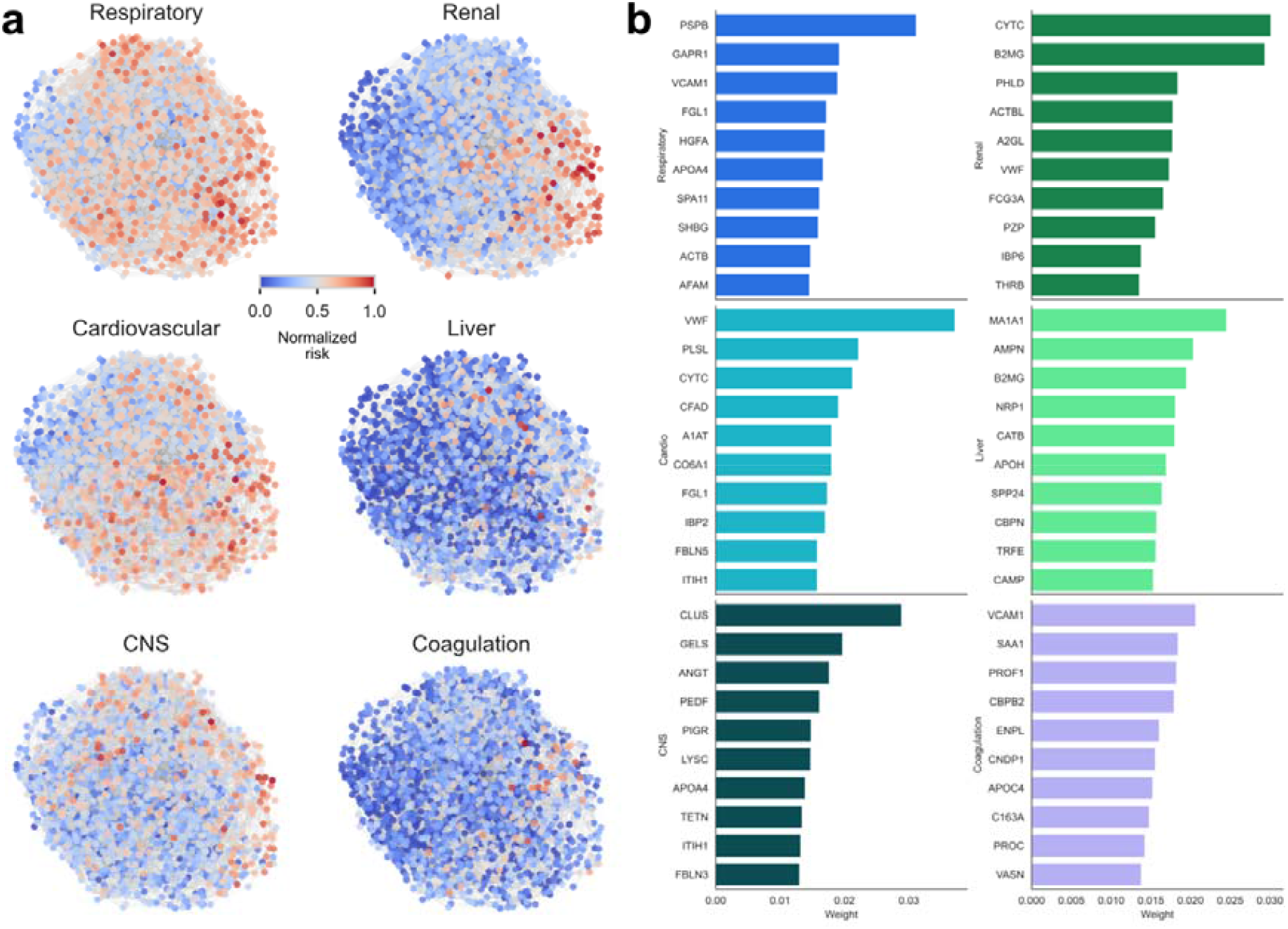
Organ dysfunction-informed digital twin maps. **a)** Projections of the organ dysfunction digital twin model colored by the local rates of organ dysfunction in the combined digital twin model built using the training, testing, and validation cohorts. The organ dysfunction values are calculated using organ specific SOFA scores for each 6 organ system. **b)** The top-10 most highly weighted proteins for the neighborhood component analysis used to construct the organ dysfunction-informed digital twin model.

## Supplementary Tables

**Supplementary Table T1a.**
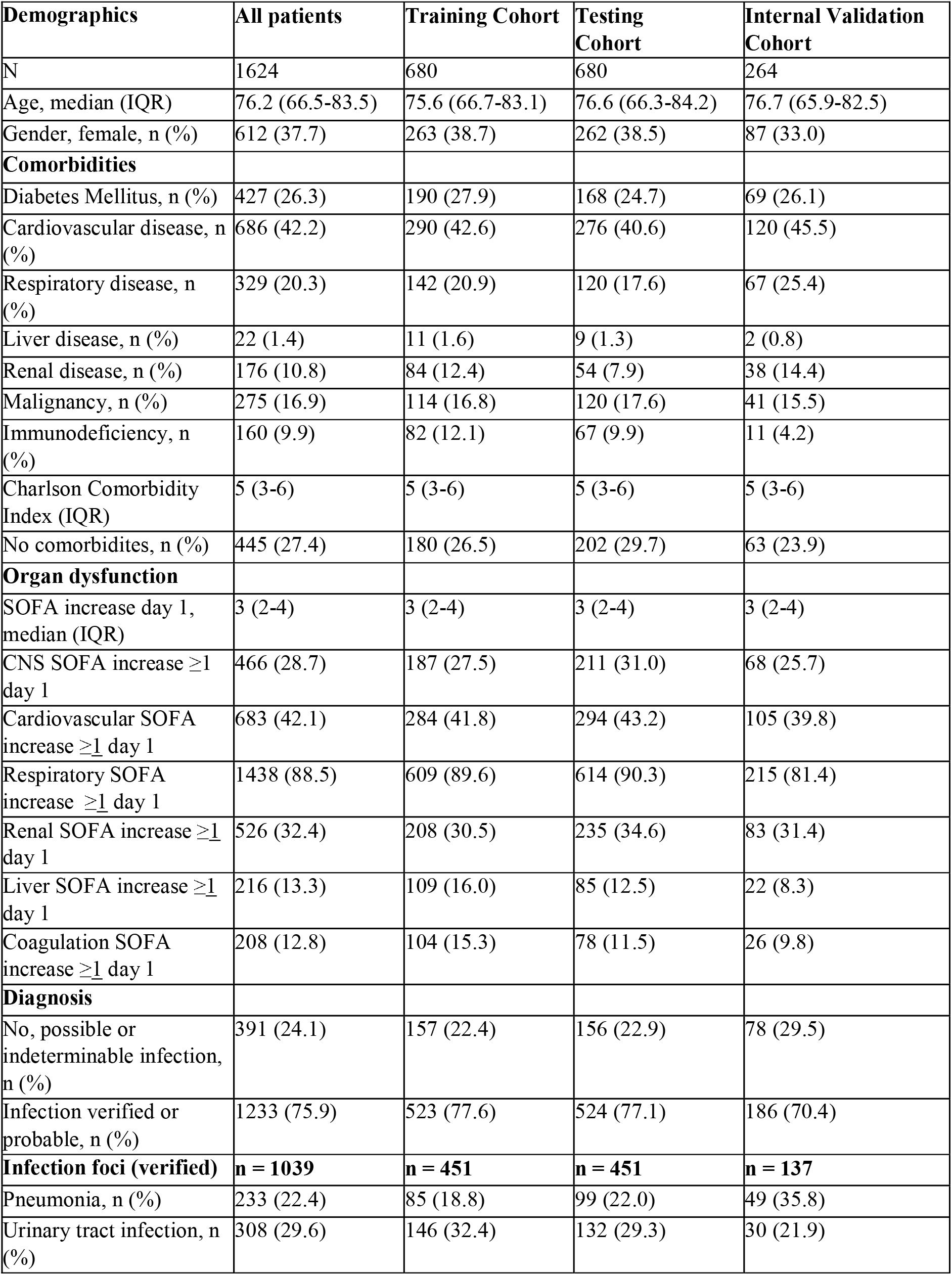
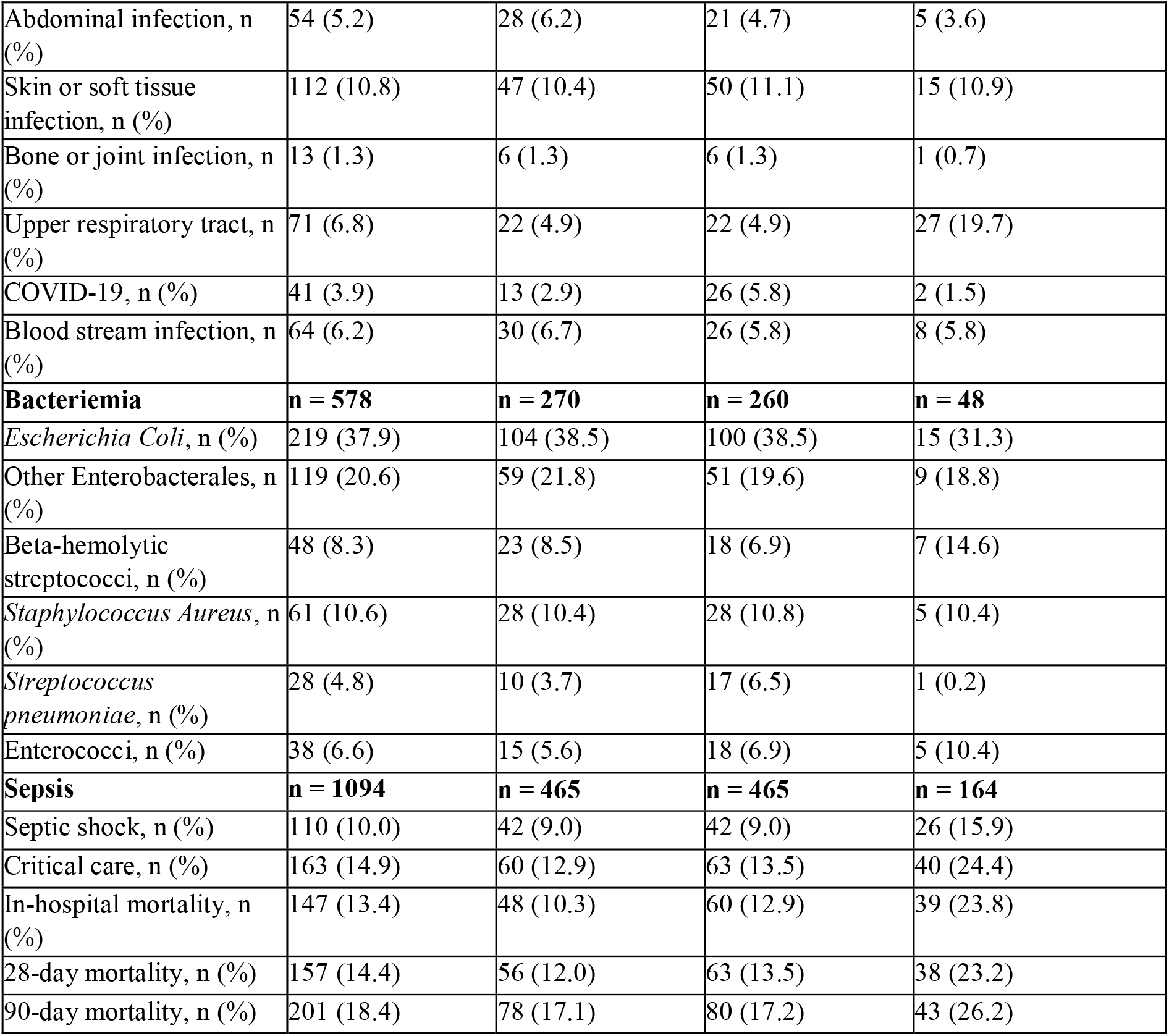
Demographic and clinical characteristics for the training, testing and validation cohorts.

**Supplementary Table T1b.**
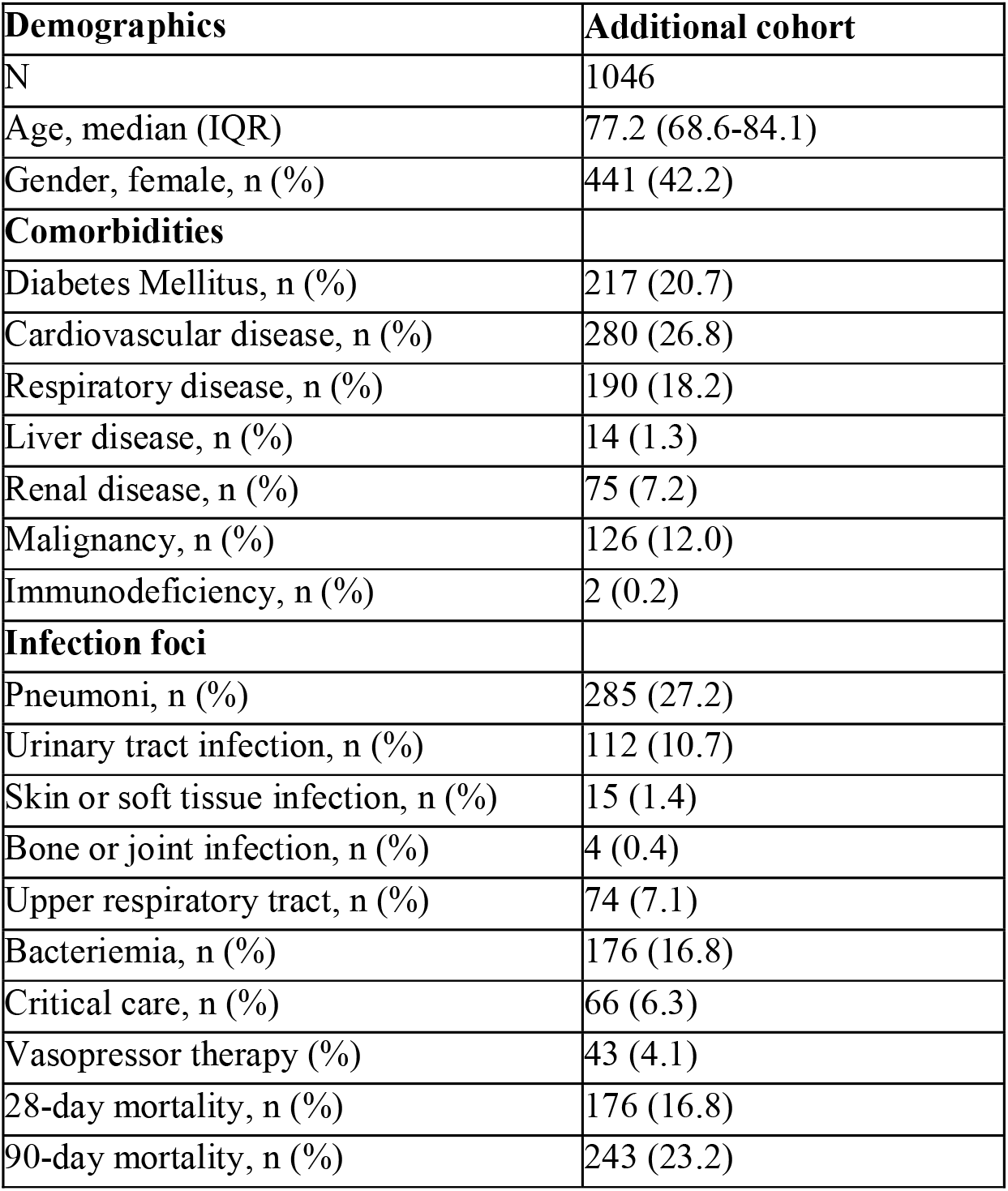
Demographic and clinical characteristics for the additional cohort.

**Supplementary Table T1c.** Demographic and clinical characteristics for the external cohort.

**Supplementary Table T1b.** Demographic and clinical characteristics for the additional cohort
| **Demographics** | **External validation** |
| --- | --- |
| N | 350 |
| Age, median (IQR) | 74.5 (62.6-83.4) |
| Gender, female, n (%) | 176 (50.3%) |
| **Comorbidities** |  |
| Diabetes Mellitus, n (%) | 65 (18.6%) |
| Cardiovascular disease, n (%) | 174 (49.7%) |
| Respiratory disease, n (%) | 94 (26.9%) |
| Liver disease, n (%) | 5 (1.4%) |
| Renal disease, n (%) | 36 (10.3%) |
| Malignancy, n (%) | 40 (11.4%) |
| Immunodeficiency, n (%) | 6 (1.7%) |
| No comorbidities, n (%) | 103 (29.4%) |
| **Diagnosis** |  |
| No, possible or indeterminable infection, n (%) | 210 (60.0%) |
| Infection probable or verified, n (%) | 140 (40.0%) |
| **Infection foci** | **n=140** |
| Respiratory infection, n (%) | 61 (43.6%) |
| Urinary tract infection, n (%) | 9 (6.4%) |
| COPD-exacerbation, n (%) | 10 (7.1) |
| Viral infection, n (%) | 32 (22.9) |
| COVID-19, n (%) | 0 |
| Bacteriemia | n=12 (8.6%) |
| **Sepsis** | **n= 89** |
| Critical care, n (%) | 3 (3.4%) |
| 28-day mortality, n (%) | 18 (20.2%) |
| 90-day mortality, n (%) | 31 (34.8%) |

**Supplementary Table T2:**
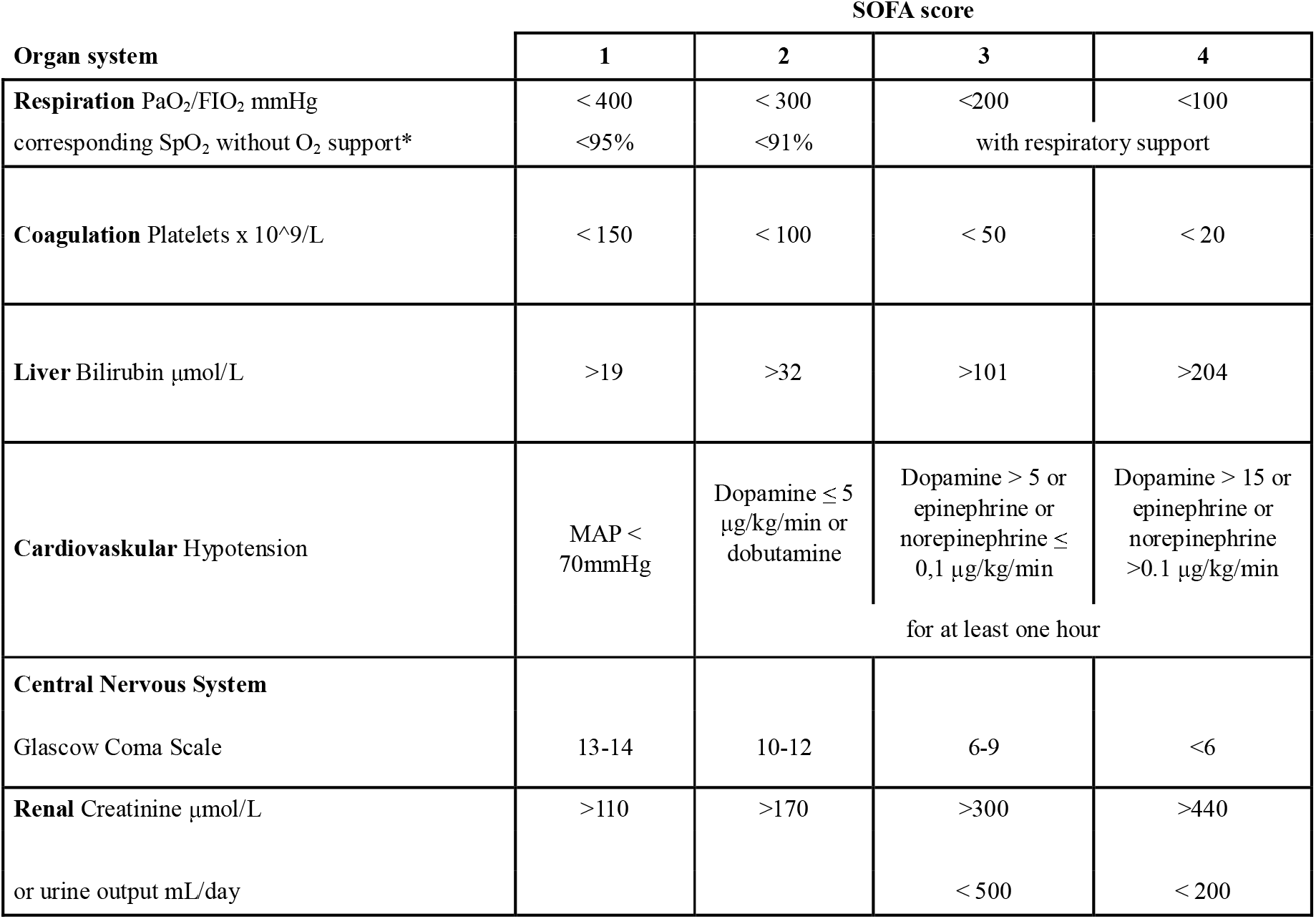
SOFA Definitions.

**Supplementary Table T3:** LC-MS/MS Acquisition windows for training and testing cohorts.

| #MS Type | Cycle ID | 1/K0 Begin [Vs/cm2] | 1/K0 End [Vs/cm2] | Start Mass [m/z] | End Mass [m/z] |
| --- | --- | --- | --- | --- | --- |
| PASEF | 1 | 0.83 | 1.6 | 681.04 | 708.33 |
| PASEF | 1 | 0.6 | 0.83 | 258.48 | 409.55 |
| PASEF | 2 | 0.88 | 1.6 | 708.33 | 733.83 |
| PASEF | 2 | 0.6 | 0.88 | 409.55 | 444.26 |
| PASEF | 3 | 0.89 | 1.6 | 733.83 | 760.32 |
| PASEF | 3 | 0.6 | 0.89 | 444.26 | 474.76 |
| PASEF | 4 | 0.9 | 1.6 | 760.32 | 790.62 |
| PASEF | 4 | 0.6 | 0.9 | 474.76 | 502.25 |
| PASEF | 5 | 0.92 | 1.6 | 790.62 | 823.07 |
| PASEF | 5 | 0.6 | 0.92 | 502.25 | 525.91 |
| PASEF | 6 | 0.93 | 1.6 | 823.07 | 858.4 |
| PASEF | 6 | 0.6 | 0.93 | 525.91 | 548.77 |
| PASEF | 7 | 0.94 | 1.6 | 858.4 | 899.09 |
| PASEF | 7 | 0.6 | 0.94 | 548.77 | 571.27 |
| PASEF | 8 | 0.96 | 1.6 | 899.09 | 948.46 |
| PASEF | 8 | 0.6 | 0.96 | 571.27 | 592.32 |
| PASEF | 9 | 0.98 | 1.6 | 948.46 | 1010.99 |
| PASEF | 9 | 0.6 | 0.98 | 592.32 | 612.8 |
| PASEF | 10 | 0.99 | 1.6 | 1010.99 | 1079.07 |
| PASEF | 10 | 0.6 | 0.99 | 612.8 | 634.8 |
| PASEF | 11 | 1.02 | 1.6 | 1079.07 | 1187.54 |
| PASEF | 11 | 0.6 | 1.02 | 634.8 | 656.97 |
| PASEF | 12 | 1.08 | 1.6 | 1187.54 | 1443.66 |
| PASEF | 12 | 0.6 | 1.08 | 656.97 | 681.04 |

**Supplementary Table T4:** LC-MS/MS acquisition windows for the validation and external cohorts.

| #MS Type | Cycle Id | Start IM [1/K0] | End IM [1/K0] | Start Mass [m/z] | End Mass [m/z] | CE [eV] |
| --- | --- | --- | --- | --- | --- | --- |
| MS1 | 0 | - | - | - | - | - |
| PASEF | 1 | 0.7000 | 0.8500 | 349.51 | 424.24 | - |
| PASEF | 1 | 0.8500 | 1.3000 | 673.34 | 700.08 | - |
| PASEF | 2 | 0.7000 | 0.8800 | 424.24 | 447.49 | - |
| PASEF | 2 | 0.8800 | 1.3000 | 700.08 | 728.00 | - |
| PASEF | 3 | 0.7000 | 0.9000 | 447.49 | 468.51 | - |
| PASEF | 3 | 0.9000 | 1.3000 | 728.00 | 758.02 | - |
| PASEF | 4 | 0.7000 | 0.9100 | 468.51 | 489.24 | - |
| PASEF | 4 | 0.9100 | 1.3000 | 758.02 | 788.71 | - |
| PASEF | 5 | 0.7000 | 0.9300 | 489.24 | 510.24 | - |
| PASEF | 5 | 0.9300 | 1.3000 | 788.71 | 821.70 | - |
| PASEF | 6 | 0.7000 | 0.9400 | 510.24 | 531.30 | - |
| PASEF | 6 | 0.9400 | 1.3000 | 821.70 | 856.42 | - |
| PASEF | 7 | 0.7000 | 0.9600 | 531.30 | 552.80 | - |
| PASEF | 7 | 0.9600 | 1.3000 | 856.42 | 893.46 | - |
| PASEF | 8 | 0.7000 | 0.9700 | 552.80 | 574.78 | - |
| PASEF | 8 | 0.9700 | 1.3000 | 893.46 | 937.11 | - |
| PASEF | 9 | 0.7000 | 0.9900 | 574.78 | 597.80 | - |
| PASEF | 9 | 0.9900 | 1.3000 | 937.11 | 985.03 | - |
| PASEF | 10 | 0.7000 | 1.0100 | 597.80 | 621.83 | - |
| PASEF | 10 | 1.0100 | 1.3000 | 985.03 | 1042.20 | - |
| PASEF | 11 | 0.7000 | 1.0400 | 621.83 | 646.84 | - |
| PASEF | 11 | 1.0400 | 1.3000 | 1042.20 | 1110.59 | - |
| PASEF | 12 | 0.7000 | 1.0700 | 646.84 | 673.34 | - |
| PASEF | 12 | 1.0700 | 1.3000 | 1110.59 | 1200.00 | - |

